# Reduced SARS-CoV-2 infection and altered antiviral transcriptional response in IBD intestinal organoids

**DOI:** 10.1101/2023.06.05.23290961

**Authors:** Barbara Jelusic, Stefan Boerno, Philipp Wurm, Nicole Przysiecki, Christina Watschinger, Stella Wolfgruber, Margit Anthofer, Sandra Ehman, Sven Klages, Kurt Zatloukal, Bernd Timmermann, Alexander Moschen, Gregor Gorkiewicz

## Abstract

IBD is characterized by altered immune reactions and infections are thought to trigger the chronic inflammatory response in IBD. The gut represents a productive reservoir for SARS-CoV-2 and the aforementioned factors together with immunosuppression used to treat IBD are likely influencing the outcomes of IBD patients in COVID-19. We used large and small intestinal organoids from IBD patients and controls to comparatively assess the transcriptional response of the gut epithelium during SARS- CoV-2 infection. Our analysis showed that IBD epithelia exhibit reduced viral loads compared to controls associated with a reduced expression of SARS-CoV-2 entry factors including the host receptor ACE2. Moreover, several genes implicated in the epithelial response to viral infection are intrinsically altered in IBD likely counteracting viral propagation. Notably, differences between IBD phenotypes exist wherein ulcerative colitis represents with induced cell death pathways and an induction of IL-1β despite overall lower viral loads suggestive of increased epithelial stress in this IBD phenotype. Altogether our analysis shows that IBD epithelia are not more prone to SARS-CoV-2 infection but epithelia from ulcerative colitis and Crohn’s disease exhibit specific differences which might explain the differing COVID-19 outcomes between IBD phenotypes.

## Introduction

Inflammatory bowel diseases (IBD), represented by Crohn’s disease (CD) and ulcerative colitis (UC), are chronic inflammatory disorders that occur in genetically susceptible individuals because of a dysregulated intestinal immune homeostasis^1^. IBD patients are prone to viral infections also driven by the immuno-suppression used to treat IBD^2, 3^. This vulnerability has raised concerns whether IBD patients represent a population at risk for worse COVID-19 outcomes. Although the incidence of COVID-19 in IBD patients is comparable to that of the general population^4, 5^, IBD patients show increased gastrointestinal (GI) symptoms during COVID-19^6^, while active IBD and corticosteroids in combination with biologics seem to increase COVID-19 severity^5–8^.

The GI tract represents a productive reservoir for SARS-CoV-2. A high expression of viral entry factors in enterocytes, like the host receptor ACE2, facilitates infection, virus amplification and faecal shedding^9–11^. Viral persistence in the GI tract likely perpetuates dissemination and pandemic spread^12, 13^. Genetic alterations in IBD can be found in genes related to immunity, and experimental models suggest that a combination of these genetic changes with viral infections drive the chronic inflammatory pathology of IBD^14–16^. In addition, the expression of key SARS-CoV-2 entry-related genes is altered in IBD and differs between ileum and colon^17–19^. Since ACE2 is considered to be tissue-protective with anti-inflammatory capabilities^20^, it’s altered expression in IBD and during SARS-CoV-2 infection might aggravate the inflammatory pathology^21, 22^. For all these reasons, it is likely that SARS-CoV-2 infection of the GI tract has profound functional consequences and might impact IBD patients differently compared to healthy persons, which might also lead to differing long term consequences. Indeed, recent data suggest that SARS-CoV-2 can persist in the GI mucosa of IBD patients over a prolonged period of time, which correlates with post-acute COVID-19 symptoms^23^.

In the current study, we used small and large intestinal organoids from IBD patients and controls to comparatively assess infection and the transcriptional response of epithelial cells as an entry site for SARS-CoV-2. We showed that IBD epithelia are generally not more prone to infection, but respond differently compared to non-IBD epithelia, which is dependent on the IBD phenotype. Importantly, intrinsic changes in the transcriptional response to virus infection seem to counteract SARS-CoV-2 in IBD, with specific differences between CD and UC.

## Results

### Modelling SARS-CoV-2 infection with small and large intestinal organoids from IBD patients and controls

Intestinal organoids were successfully cultivated from 6 patients with CD (CD ileum), 5 patients with UC (UC colon), as well as 6 healthy ileum (h-ileum) and 5 healthy colon (h-colon) controls (Fig. 1). 3D organoids and 2D organoid monolayers were generated in parallel, the latter used to model a luminal SARS-CoV-2 infection. Infection was performed with the SARS-CoV-2 strain hCoV-19/Austria/Graz-MUG5 (B.1.5/O/20A) isolated from a COVID-19 patient from the first pandemic wave^24^. 2D organoid monolayers were infected at an MOI of 0.1 and collected 48 hours post-infection (p.i.), and 3D organoids were infected at an MOI of 1 and collected 72 hours p.i. based on previous protocols^25, 26^. Experimental set-up consisted of four organoid monolayers per patient, two infected and two uninfected, as well as duplicates of 3D organoids per patient, one infected and the other uninfected. RNA-seq generated from organoid monolayers yielded on average 29 million reads per sample (single monolayer; SD = 3.7 M). High correlation of RNA-seq reads from monolayer duplicates allowed for their pooling in downstream biostatistical analyses (Suppl. Fig. 1).

**Figure 1.**
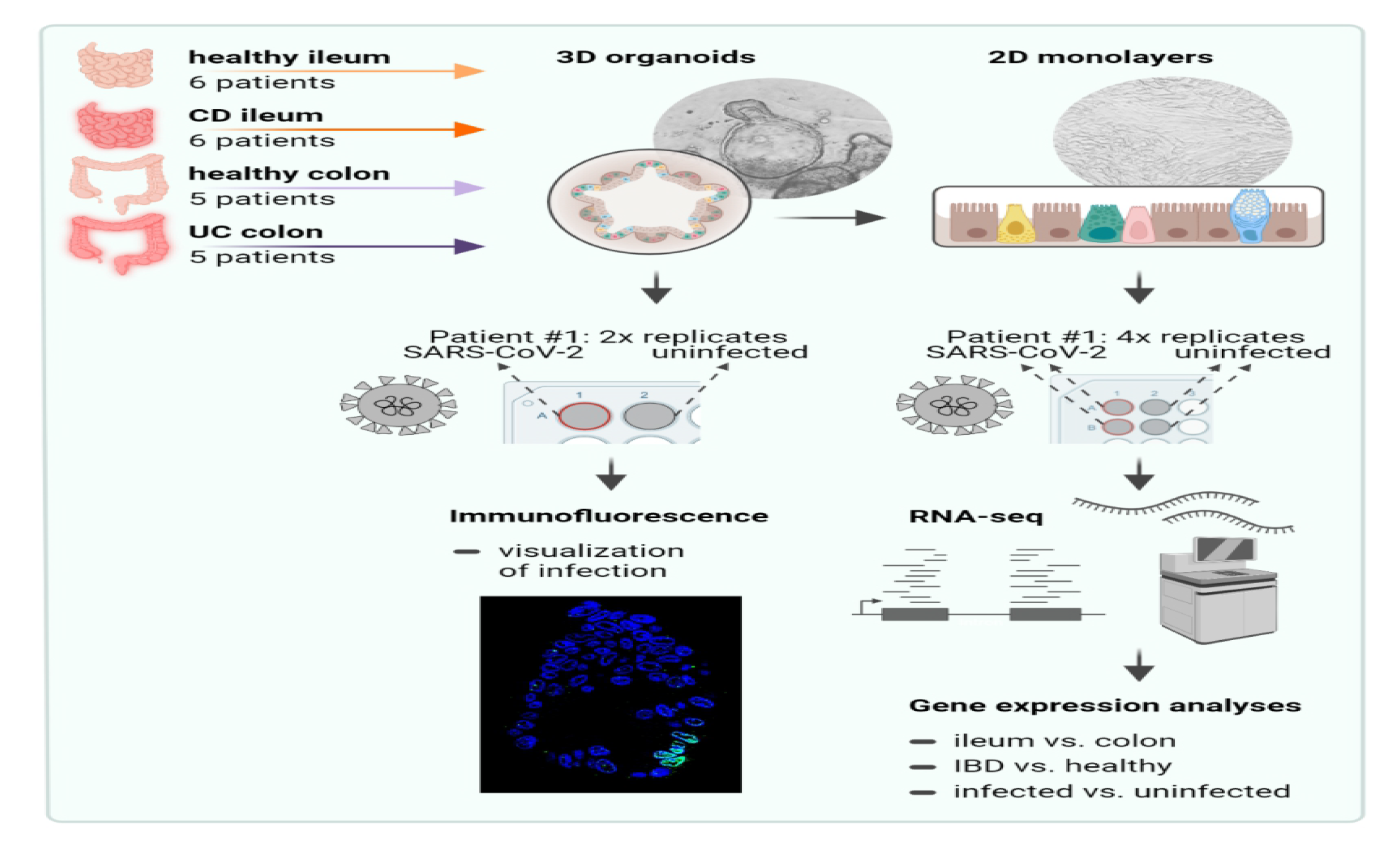
Modelling SARS-CoV-2 infection with small and large intestinal organoids from IBD patients and controls. 3D organoids were cultured from biopsy specimens retrieved from endoscopy of CD patients (ileum), UC patients (colon) and healthy controls (ileum and colon). Four 2D monolayer replicates were created from the 3D organoids of each patient. Two replicates were inoculated with SARS-CoV-2. Samples from monolayers were collected after 48 hours and subjected to RNA sequencing. Differential gene expression analyses between ileum vs. colon, IBD vs. healthy and infected vs. uninfected samples were performed from each patient. Remaining 3D organoids were infected and collected after 72 hours to visualize infection by immunofluorescence.

### Tissue origin followed by IBD status but not SARS-CoV-2 infection are the major discriminators in deep transcriptomic analyses

Principal component analysis (PCA) based on gene counts separated organoid-derived monolayers most according to tissue origin (Fig. 2A). H-ileum clearly separated from h-colon and UC colon (PC1, 36.3 % variation) while CD ileum samples were more dispersed and more similar to colon samples (Fig. 2A, Suppl. Fig. 2B). SARS-CoV-2 infection induced only small variations in PCA. The major separators in PCA were genes differentiating small and large intestine, for example, the colon marker *MUC12* or the small intestinal marker *APOB* (Fig. 2B). IBD samples were separated from healthy controls mostly by PC4 (5.61 % variation) driven by differential gene expression of *UGT2B17*, *APOA4, CEACAM7* and *NXPE1*, known to be altered in IBD^15, 28, 29^. Organoids retained typical expression signatures of their original tissues including markers of stemness, enterocytes, Paneth cells, enteroendocrine cells and goblet cells, also evident in IBD organoids (Fig. 2C). Interestingly, three CD ileum cases clustered together with colon samples, suggesting that CD can alter gene expression to a more colon-like phenotype in at least certain individuals^30^. Of note, the level of mitochondrial transcripts used as a marker for cell damage was not different between the organoid groups^31^ (Suppl. Fig. 2C).

**Figure 2.**
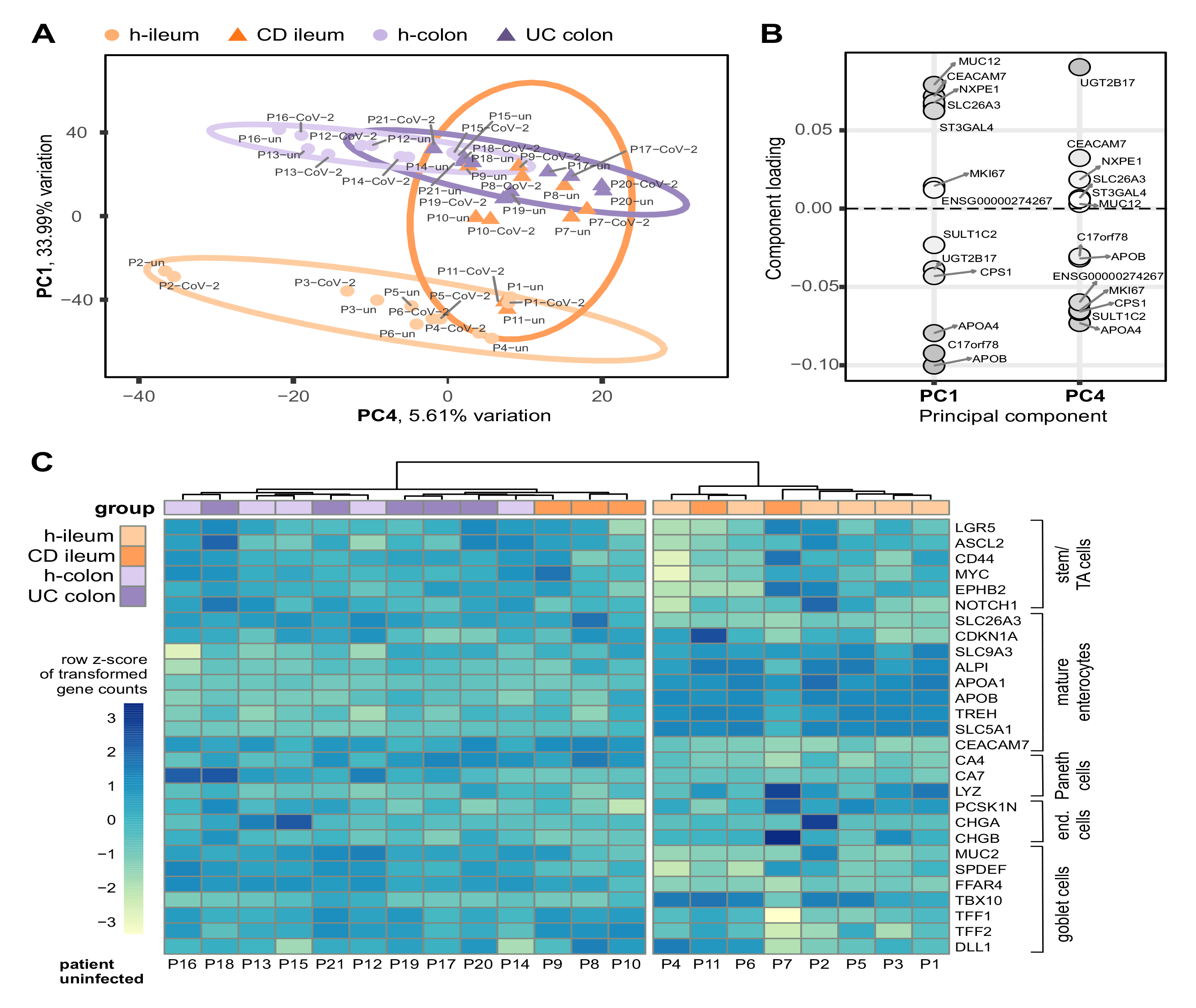
Tissue origin followed by IBD status but not SARS-CoV-2 infection are the major discriminators in deep transcriptomic analyses. **A)** Principal component analysis (PCA). Sample distances were calculated from transformed normalized gene counts. Ellipses indicate 95% confidence intervals around sample groups. Each dot represents pooled organoid monolayer replicates from one patient, either treated with SARS-COV-2 or non-infected (ni). **B)** Genes that are top 10% contributors to variation along PC1 and PC4. A darker grey marks a stronger influence on a PC. **C)** Heatmap with sample clustering showing cell type marker genes for small and large intestines. Marker genes are stratified into stem/transit-amplifying cells (TA), mature enterocytes, Paneth cells, enteroendocrine cells and goblet cells. Row Z-Score is calculated based on transformed normalized counts. N(patients) = 5-6; CD = Crohn’s disease; UC = ulcerative colitis.

### SARS-CoV-2 infection is highest in healthy ileum while IBD status does not increase viral load

3D organoids from all infected organoid groups stained with an antibody against the SARS-CoV-2 spike (S) protein showed intra-cytoplasmic signals often gathering around the nucleus, which were absent in uninfected controls (Fig. 3A, Suppl. Fig. 2A). 16.76 viral gene counts (median; IQR = 11.23 - 26.40) compared to 14.75 × 10^6^ human gene counts (median; IQR = 14.33 × 10^6^- 15.60 × 10^6^) were generated per infected single organoid-derived monolayer. Viral transcripts were evenly distributed along the SARS-CoV-2 genome (Fig. 3B). We hypothesized that the viral load, defined as the sum of all normalized viral gene counts per patient, would be greater in ileum than in colon due to the known higher *ACE2* expression in ileum^19^. Indeed, h-ileum showed the highest viral load compared to other infected organoid groups (Fig. 3A). CD ileum showed significantly less viral transcripts than h-ileum and there was also a trend towards reduced viral load in UC colon compared to h-colon. SARS-CoV-2 cell entry is facilitated by the host receptor ACE2 and proteases CTSL, CTSB, furin, TMPRSS2/4/11D/13 and NRP1^32–37^. *ACE2* expression was highest in h-ileum (Fig. 3D) and expression in organoids mirrored the known decrease in the inflamed ileum of IBD patients as well as differences between ileum and colon^19, 38^. Conversely, *ACE2* expression is reported to be increased in the colon upon inflammation^38^, which corresponds to a non-significant trend of increased *ACE2* expression in UC colon organoids compared to h-colon. *TMPRSS2* expression, the major protease facilitating viral cell surface entry^33^, was significantly increased in CD ileum compared to h-ileum corroborating findings from CD patients^39^. Expression of *CTSL* and *CTSB*, both proteases important for endosomal entry of the virus^33, 40^, followed the same trend as *ACE2*. *TMPRSS4*, *FURIN* and *NRP1* were similarly expressed across all groups, while *TMPRSS11D* was not detectable (Suppl. Fig. 3). Notably, SARS-CoV-2 infection did not significantly alter expression of any investigated entry gene in our model (Fig. 3D, Suppl. Fig. 3). Correlation analysis of viral load and the expression of entry genes revealed a significant positive correlation of *ACE2*, *CTSL* and *CTSB* with viral load, but only in organoids from controls and not IBD cases, suggesting that SARS-CoV-2 entry and propagation mechanisms might be altered in IBD (Fig. 3E, Suppl. Fig. 4). In summary, our organoid model recapitulated the expression of SARS-CoV-2 entry genes in small and large intestines and showed changes typically occurring in IBD. However, although the IBD phenotype modulates the expression of entry genes, it does not increase SARS-CoV- 2 infection levels, which were even lower in IBD organoids.

**Figure 3.**
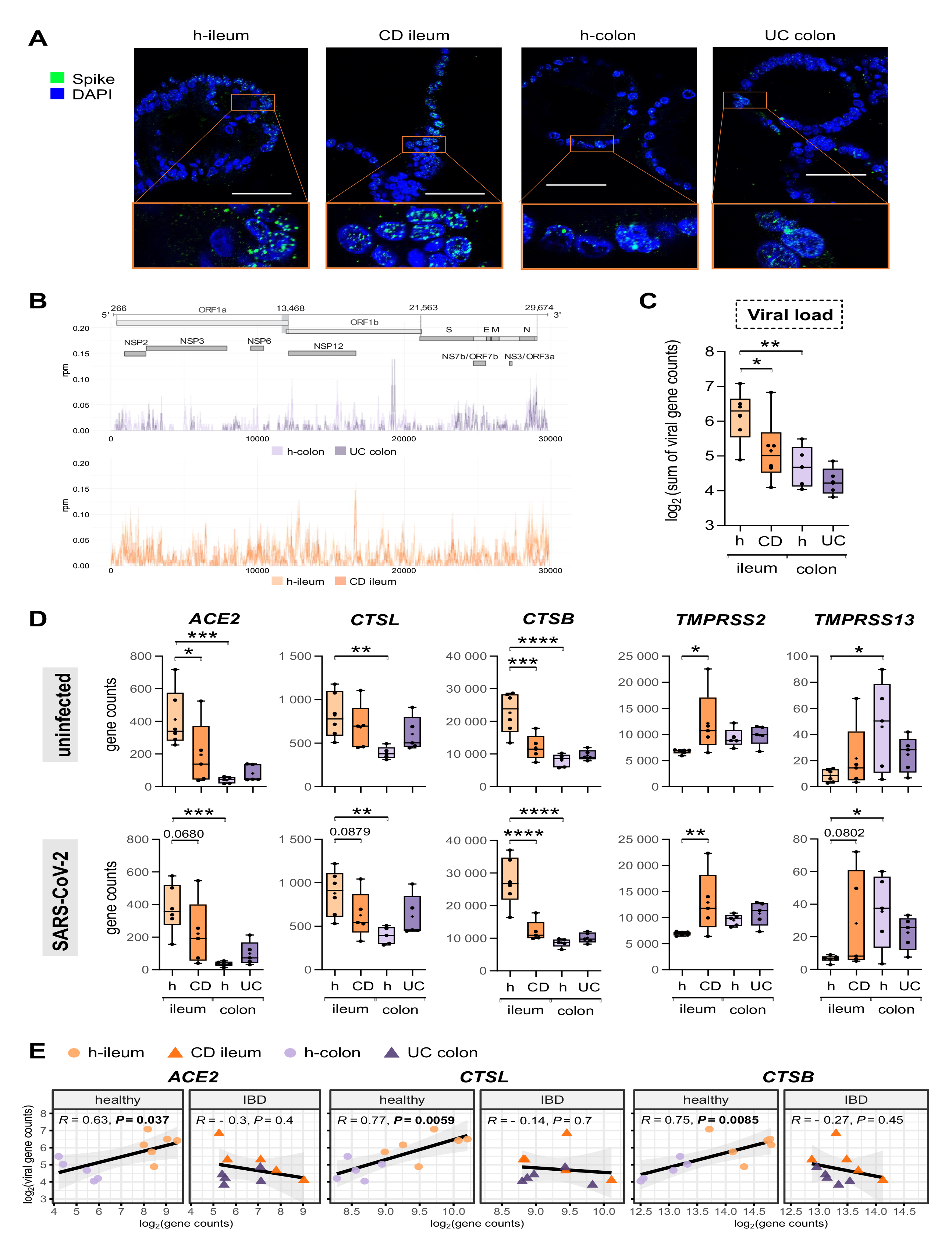
SARS-CoV-2 infection is highest in healthy ileum while IBD status does not increase viral load. **A)** Representative immunofluorescence staining of the SARS-CoV-2 spike (S) protein in 3D organoids (green). DAPI counterstain (blue). Bar represents 50 μm. **B)** Distribution of viral reads along the SARS-CoV-2 genome. Cumulative coverage of plus and minus strand transcripts is shown (median in bold). **C)** Viral load in 2D organoid-based monolayers 48 hours post infection determined by viral transcripts normalized by reads per million (rpm) and summed per sample (mean ± SD). **D)** SARS-CoV-2 entry gene expression in uninfected and infected 2D organoid groups (mean ± SD; 1way ANOVA with uncorrected Fisher’s LSD on following comparisons: h-ileum vs. CD ileum, h-ileum vs. h-colon, h-colon vs. UC colon. **E)** Viral load in 2D cultures correlates to SARS-CoV-2 entry genes only in non-IBD groups (Pearson correlation of transformed normalized gene counts). All panels: each dot represents one patient, pooled monolayer replicates. N(patients) = 5-6; CD = Crohn’s disease; UC = ulcerative colitis, * *P* < 0.05, ** *P* <0.01, *** *P* < 0.001, **** *P* < 0.0001.

### SARS-CoV-2 infection impacts gene expression differently depending on intestinal origin and IBD status of organoids

Our study design allowed for the comparison of paired infected and uninfected organoids originating from the same individual reducing the influence of inter-patient variation (Fig. 1A). 995 differentially expressed genes (DEGs, *P* < 0.05) were found in h-ileum organoids upon infection, 802 DEGs in CD ileum, 760 in h-colon and 895 in UC colon (Suppl. File 1). H-ileum showed most unique DEGs consistent with the dominant infection of this organoid group (Fig. 4A, Suppl. Fig. 5A). Organoids from healthy controls shared more DEGs than IBD organoids upon infection, indicating a greater inter-individual variation in IBD (Suppl. Fig. 5B-C). Over-representation pathway analysis (ORA; Reactome^41^) revealed altered pathways of immunity, central cellular functions and signalling cascades upon infection. Surprisingly, enriched pathways were often shared between h-ileum and UC colon while depleted pathways seemed more specific for tissue type and IBD status (Fig 4B, Suppl. File 2). These findings were mirrored in the gene set enrichment analysis (GSEA; Reactome; Suppl. Fig. 6, Suppl. File 3). Grouping these identified pathways into biological themes showed an enrichment of extracellular matrix organization, cell death, metabolism, immunity and signalling cascades upon infection (Fig. 4C). Amongst these were FOXO-mediated transcription, TGF-β, EGF, MAPK and NOTCH signalling, which are implicated in the inflammatory response and known to be altered by coronavirus infection^42–46^ (Suppl. File 3). Circadian clock-related genes showed a consistent and strong upregulation in all organoid groups, a process which controls host-virus interactions and which can be manipulated by viruses^47^. Cell death pathways, including apoptosis and necrosis, were stronger induced in h-ileum compared to CD ileum, consistent with the increased infection level in this organoid group (Fig. 4D). However, cell death was also stronger induced in UC colon compared to h-colon despite a lower infection level, suggesting that SARS-CoV-2 leads to a stronger transcriptional induction of cell death in UC. Coherent to this finding was also a difference in defence signalling pathways. A vital defence mechanism used by epithelial cells against viruses, the interferon type I (alpha/beta) signalling, was induced in UC colon but not in h-colon (Fig. 4D). Taken together, SARS-CoV-2 infection influenced gene expression of GI organoids differently, depending on tissue origin and IBD status. H-ileum and UC colon showed many commonalities and UC colon seemed to react with increased epithelial stress upon SARS-CoV-2 infection.

**Figure 4.**
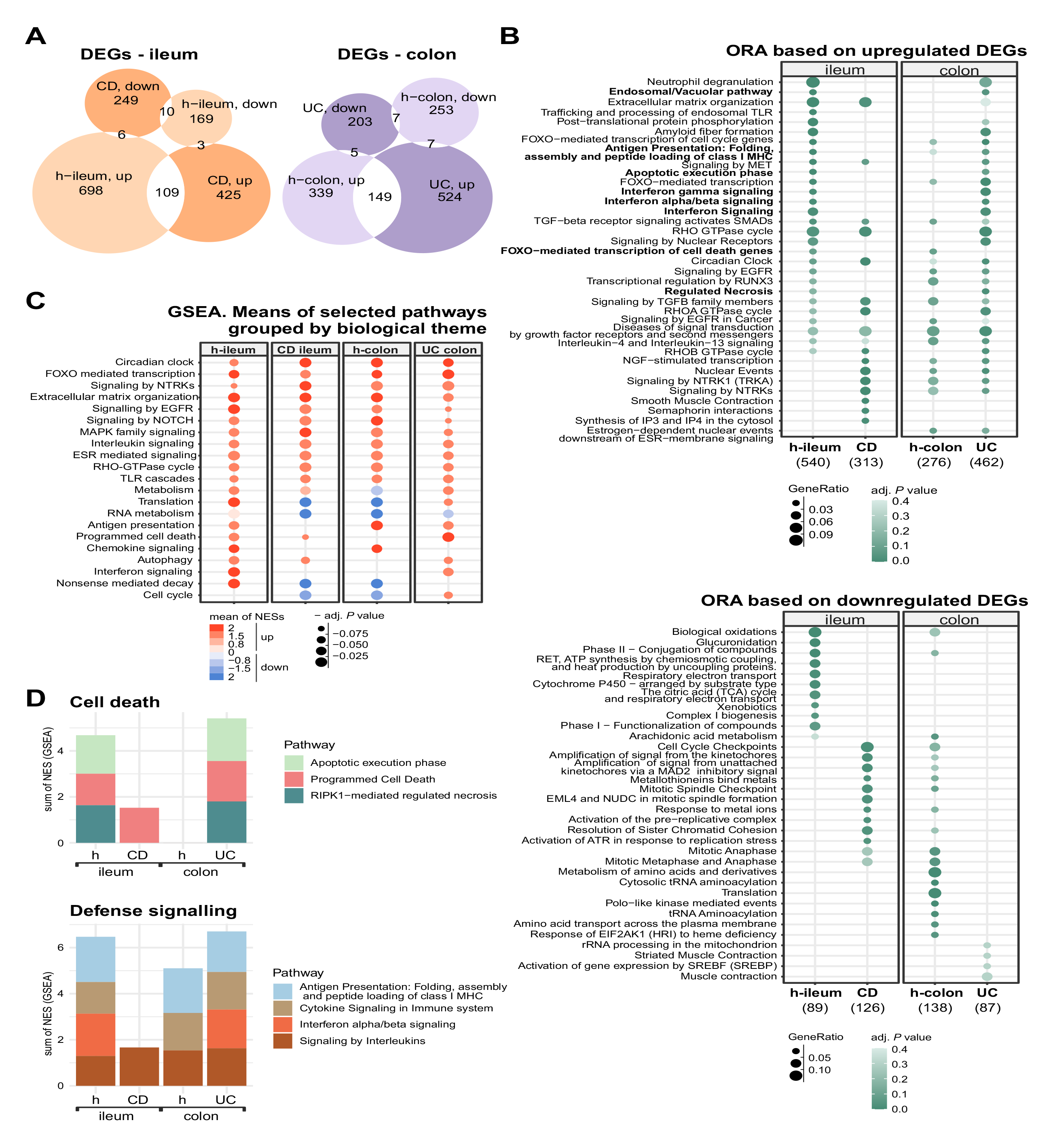
SARS-CoV-2 infection impacts gene expression differently depending on intestinal origin and IBD status of organoids. **A)** Venn diagram specifying upregulated (up) and downregulated (down) DEGs (*P* < 0.05) in SARS-CoV-2-infected 2D organoid-based monolayers compared to uninfected controls. **B)** Over-representation analysis (ORA; Reactome) of most significantly changed pathways based on up- (left) and downregulated (right) DEGs upon infection. **C)** Gene set enrichment analysis (GSEA; Reactome) based on sign of log2-fold change and *P* values of all detected genes upon infection. Results (Suppl. File 3) were combined for simplicity. Pathways were selected based on their prevalence across groups or relevance to viral infections before grouping into common biological themes. **D)** Apoptosis, necrosis and interferon alpha/beta signalling were enriched only in h-ileum and UC colon upon infection. Normalized enrichment scores determined by GSEA for chosen enriched pathways are stacked for each group (note: some genes overlap in some gene sets). Statistics of differential expression were calculated with the Wald test. N(patients) = 5-6, CD = Crohn’s disease, UC = ulcerative colitis.

### Expression of genes implicated in the cellular response to viral infection is intrinsically altered in IBD

IBD is characterized by altered expression of genes implicated in immunity^48^. These alterations are central to the pathogenesis of IBD but might also influence the cellular reaction and susceptibility to infections. Comparison of uninfected IBD organoids to uninfected controls revealed 3232 DEGs (*P* < 0.05) in CD ileum and 2775 DEGs in UC colon (Suppl. Fig. 7, Suppl. File 4). We next focused our analysis to genes implicated in the host defence against viruses, including the Gene Ontology gene sets ‘response to virus’ (GO:0009615) and ‘type I interferon signalling pathway’ (GO:0060337) with type III interferons and their receptors amounting 330 genes in total (Fig. 5A, Suppl. File 5). Of these genes, 44 were differentially expressed in CD ileum compared to h-ileum and 29 genes were differentially expressed in UC colon compared to h-colon (Fig. 5B, Suppl. File 5). These genes could be functionally categorized into interferon-stimulated genes (ISGs), factors for recognition of viral RNA or with direct antiviral activity, factors regulating cellular defence mechanisms, cytokines, chemokines and their receptors, as well as antigen presentation, autophagy and cell death (Fig. 5C, Suppl. File 5). Overall, we identified 98 of these response to virus genes significantly different in the four organoid groups with infection (discussed below) or without infection. Comparing uninfected IBD organoids to controls revealed the viral RNA sensors *OASL* and *OAS1*, which degrade viral RNA and activate the retinoic acid-inducible gene I (RIG-I) leading to a type I interferon (IFN) response^49^, significantly upregulated in CD ileum. *OAS1* was shown to be protective against severe COVID-19^50^. *BST2*, a potent anti-SARS-CoV-2 factor inhibiting budding of virus particles from the cell surface^51^, and *SEC14L1* which impairs RIG-I signalling^52^ were also upregulated in CD ileum. Most significantly downregulated in uninfected CD ileum were *TKFC* (syn.: DAK), a negative regulator of antiviral signalling^53^, and the heparan sulphate biosynthesis gene *EXT1,* which represents an ancillary SARS-CoV-2 entry factor^54^. Together, 5 out of 6 top changed response to virus genes in uninfected CD ileum compared to h-ileum likely impair SARS-CoV-2 infection (Fig. 5C, 6A, Suppl. File 5). In uninfected UC colon compared to h-colon, we found the interferon regulatory transcription factor *IRF7* and the antiviral and pro-apoptotic factor *XAF1*^55^ strongly upregulated. *IRF7* is crucial in propagating signalling cascades initiated by viral infection and normally shows low constitutive expression in enterocytes^19, 56^. Interestingly, *MX2*, an ISG with antiviral activity^57^ and reported to be induced in COVID-19^58^, was also upregulated in UC colon but downregulated in CD ileum, indicating subtle differences between IBD phenotypes. Significantly downregulated in uninfected UC colon was the apoptosis regulator *BCL2,* known to be colitis-protective and whose viral homologs propagate infection by inhibiting premature apoptosis of infected cells^59, 60^. Notably, BCL2 inhibition is thought to counteract SARS-CoV-2 infection^61^. Also downregulated was *DDIT4,* which inhibits virus propagation by suppressing mTOR activity^62, 63^. Again, 4 out of 5 top changed response to virus genes likely counteract SARS-CoV-2 infection in baseline UC colon (Fig. 5C, 6B, Suppl. File 5). In summary, several genes with antiviral action are already induced in IBD organoids prior to infection. This goes in accordance with our finding that viral load was not increased in IBD organoids compared to controls. However, the expression of these response to virus-related genes are different between IBD phenotypes.

**Figure 5.**
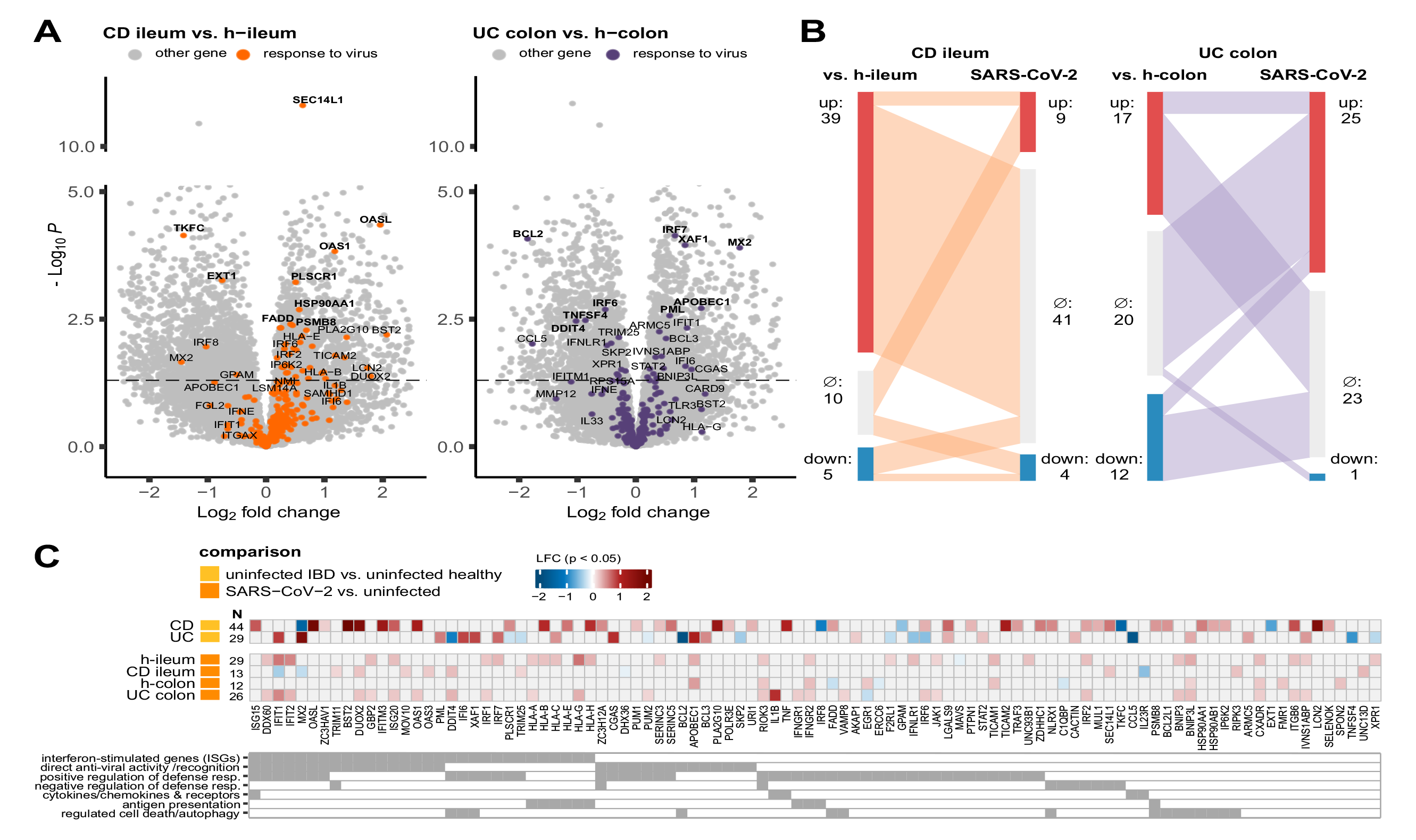
Expression of genes implicated in the cellular response to viral infection is intrinsically altered in IBD, different between ileum and colon and influenced by IBD status upon SARS-CoV-2 challenge. **A)** Volcano plot showing DEGs of uninfected CD ileum vs. healthy ileum (left) and UC colon vs. healthy colon (right). Orange and purple dots represent response to virus-related genes; horizontal dashed line marks *P* value of 0.05. Bolded genes remained significant after BH correction with FDR = 0.1. **B)** Sankey plot visualizing the amount of upregulated and downregulated DEGs in baseline IBD vs. healthy compared to infected vs. uninfected in CD ileum and UC colon. **C)** DEGs (*P* < 0.05) from the created gene set ‘response to virus’ in (yellow) IBD organoids vs. non-IBD/healthy organoids and in (orange) SARS-CoV-2 infected organoids vs. uninfected. Heatmap shows log_2_fold changes (LFC) with *P* < 0.05. Number (N) of significantly different genes per comparison is shown. Statistics of differential expression were calculated with the Wald test. N(patients) = 5-6, CD = Crohn’s disease, UC = ulcerative colitis, GOI = gene of interest.

### Expression of genes implicated in the response to viral infection are different between ileum and colon and influenced by IBD status upon SARS-CoV-2 challenge

We next assessed the effect of SARS-CoV-2 infection on the transcriptional response to virus in organoids (Suppl. Fig. 8, Suppl. File 5). Infection significantly changed 29 of these genes in h-ileum, 13 in CD ileum, 12 in h-colon and 26 in UC colon (Fig. 5C; *P* < 0.05). In CD ileum, 9 were up-and 4 downregulated and 3 of them were already altered in uninfected CD ileum. In UC colon, 25 were up-and 1 downregulated and 5 of them were already altered in uninfected UC colon (Fig. 5B-C). Similar to findings from pathway analyses, h-ileum and UC colon showed a stronger antiviral response compared to CD ileum or h-colon. Among top induced genes upon infection were *IFIT1* and *IFIT2*, known to sequester 5’PPP-capped viral RNAs in host cells, thereby inhibiting virus amplification^64, 65^. Both genes were upregulated in h-ileum and UC colon upon infection. *IFIT1* was already induced in baseline UC colon but was repressed in CD ileum upon infection. *IFIT2* was one of the strongest induced genes in h-ileum upon infection, in addition to the pro-apoptotic mitophagy gene *BNIP3L* (Fig. 5C, Fig. 6C-D). The prominent activation of *IFIT* genes suggests that these viral RNA sensing and degrading factors are central in the intestinal response to SARS-CoV-2 infection. The viral RNA-degrading exonuclease *ISG20* (induced in baseline CD ileum) was induced in h-ileum and UC colon upon infection. Induced in h-ileum but not in CD ileum upon infection were *ITGB6,* a possible alternative receptor for SARS-CoV-2^66^ and *SERINC3,* a known antiviral defence factor and a suggested risk gene for severe COVID-19^67^. Antigen presentation also seemed be modified in IBD organoids. The HLA class I genes *HLA-B,* a known IBD risk gene^68^, and *HLA-H,* were induced in CD ileum but could not be further induced upon infection in this organoid group in contrast to h-ileum. Conversely, these genes were inducible in UC colon upon infection but not in h-colon (Fig. 5C, 6C-D). The antiviral oxidase *DUOX2* and *SEC14L1,* a negative regulator of antiviral signalling and interferon production^69^, were further induced in CD ileum upon infection and the antiviral GTPase *MX2* (repressed in uninfected CD ileum) was further repressed upon infection. Strongest upregulation in UC colon upon infection showed *IL1*Β, in addition to *IFIT1* and *DDIT4.* IL-1 is a cytokine that induces IGSs and is insensitive to viral immune escape strategies^70^. As opposed to its beneficial effects against infection, it was recently shown to drive chronic intestinal inflammation in UC^71, 72^. Several response to virus genes were upregulated in both UC colon and h-colon upon infection (*RIOK3*, a negative regulator of RIG-I, *EGR1*, *BNIP3L*, *IFNGR2* and *CXADR*). Certain genes were found to be induced upon infection only in UC colon but not h-colon, and at the same time were already different in uninfected UC, such as *IFIT1*, *IVNS1ABP* (up in uninfected UC colon)*, DDIT4, PUM2* and *IRF6* (down in uninfected UC colon). The cytidine deaminase *APOBEC1*, known to inhibit viral replication^73^, was increased in baseline UC and showed the strongest induction in h-colon upon infection.

**Figure 6.**
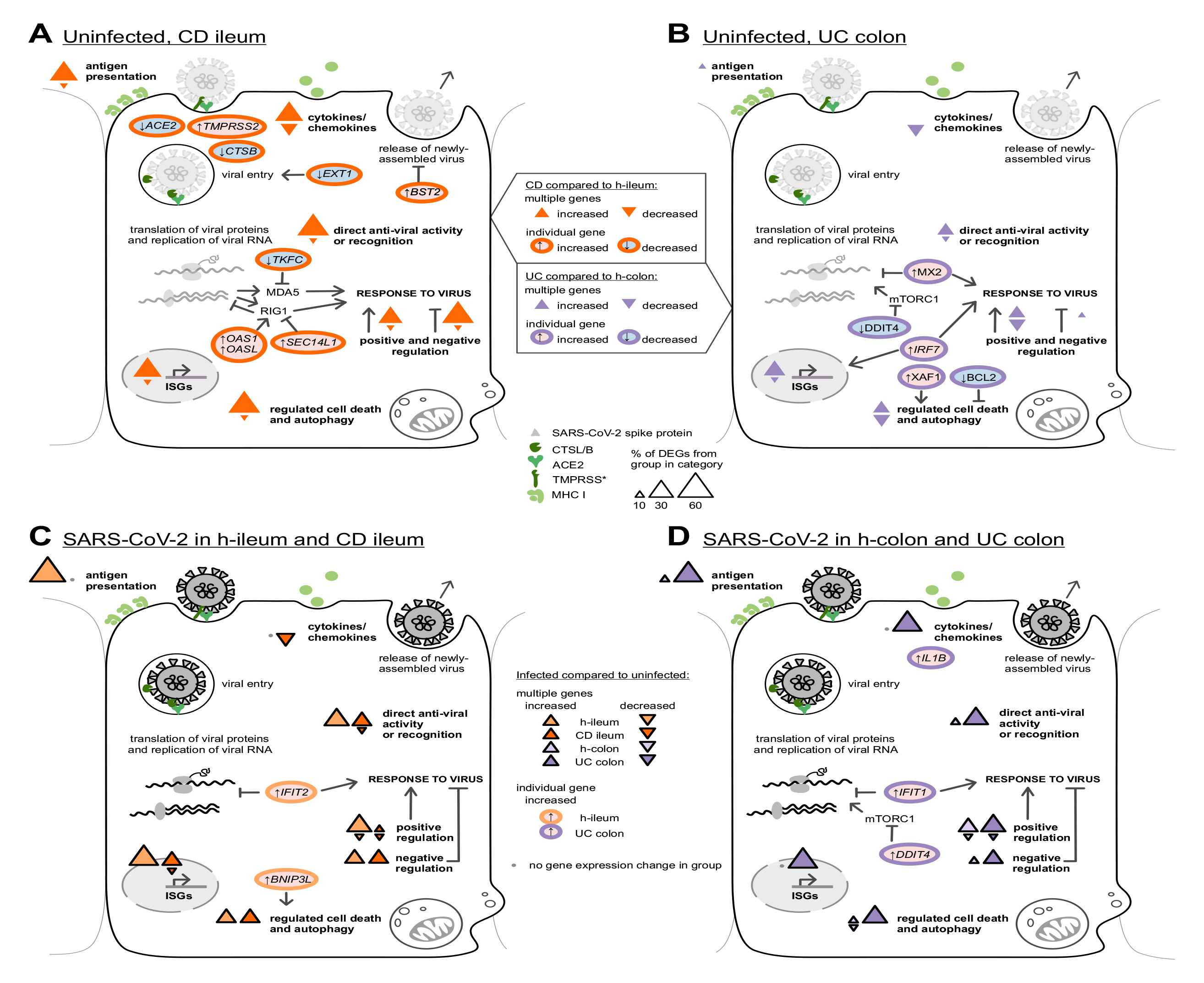
Proposed influences of the differential response to virus-related gene expression in uninfected IBD epithelia and upon SARS-CoV-2 infection. DEGs (*P* < 0.05) from the created gene set ‘response to virus’ categorized into genes important for antigen presentation, anti-viral activity or recognition, cell death, cytokine/chemokines as well as SARS-CoV-2 entry factors. **A)** Uninfected CD ileum organoids compared to uninfected h-ileum organoids. **B)** Uninfected UC colon organoids compared to uninfected h-colon organoids. **C)** SARS-CoV-2 infection in h-ileum and CD ileum. **D)** SARS-CoV-2 infection in h-colon and UC colon. Sizes of triangles correspond to the percentage of DEGs that came from a particular organoid group and comparison, i.e., which percent of DEGs in that category are changed in a particular group (see also Fig. 5C). Categories contain different number of DEGs. Chosen DEGs (*Padj* < 0.1) and SARS-CoV-2 entry genes are additionally individually depicted in ellipses. Statistics of differential expression were calculated with the Wald test. N(patients) = 5-6; CD = Crohn’s disease; UC = ulcerative colitis.

In summary, our comparative analyses showed that the transcriptional response of small and large intestinal epithelia to SARS-CoV-2 infection is altered in IBD. Figure 6 summarizes the main transcriptional alterations relevant to infection. In CD ileum organoids, genes implicated in antigen presentation, direct antiviral activity, cytokine/chemokine signalling as well as regulated cell death and autophagy were already upregulated prior to infection. In UC colon organoids compared to healthy colon, such an induction is less evident. Upon infection, h-ileum organoids react stronger and/or preserve their transcriptional response to virus to a greater extent compared to CD ileum organoids. This difference possibly originates from the higher viral load found in h-ileum. However, UC colon organoids respond stronger upon SARS-CoV-2 challenge despite a slightly reduced viral load compared to h-colon. This reaction includes cell death and autophagy but also a prominent induction of *IL1*Β expression, which is intriguing since this cytokine seems to drive inflammation in UC^71^ and COVID-19 outcomes in UC patients seem to be worse compared to CD^74^.

## Discussion

The GI tract represents a niche for productive SARS-CoV-2 infection. IBD is characterized by altered immune reactions and, therefore, it is likely that SARS-CoV-2 impacts the GI tract of IBD patients differently compared to healthy individuals. In addition, genetic differences between CD and UC might differentially impact the response to SARS-CoV-2. We used small and large intestinal organoids from IBD patients and controls to comparatively assess susceptibility and transcriptional responses to SARS-CoV-2 infection. We showed that IBD phenotype does not increase SARS-CoV-2 infection levels, which is likely associated with a depressed expression of SARS-CoV-2 entry factors (e.g. *ACE2* and *CTSL/B*) most prominent in CD ileum, but also with an upregulation of several antiviral defence genes potentially counteracting viral propagation in IBD. However, transcriptional differences between IBD phenotypes in response to SARS-CoV-2 exist, which might lead to varying disease outcomes between CD and UC.

ACE2, the host receptor of SARS-CoV-2^75^, is expressed apically in enterocytes^32^ indicating that virus attachment and uptake via the lumen is a dominant infection route in the GI tract. This was modelled in our 2D organoids^76, 77^. SARS-CoV-2 entry factors are reported to be altered in IBD ileum^17, 38, 78–80^ and we recapitulated a reduced *ACE2* expression in CD ileum organoids compared to h-ileum, which correlates with significantly reduced infection levels. In colon, *ACE2* expression increases with the inflammatory tone^17, 38, 78–81^ and we also noted a weak increase of *ACE2* expression in UC colon compared to h-colon. Despite that, the lowest infection levels were noted in this organoid group. The proteases TMPRSS2 and CTSL/B, which prime the S protein to facilitate binding to ACE2^40, 75^, are also reported to be altered in IBD^17, 18, 38, 79, 82^ and their expression was mostly recapitulated in our organoids. Entry genes were not transcriptionally changed by SARS-CoV-2 infection in our model, which is different to some other studies^83, 84^. Importantly, infection levels were not increased in our IBD organoids. We noticed a positive correlation of viral load with the expression of entry factors *ACE2* and *CTSL/B* only in non-IBD organoids, indicating that infection levels and entry factors are functionally linked in healthy epithelia but this relation is likely skewed in IBD.

We cultured organoids from multiple IBD and non-IBD individuals which allowed us to at least partially capture the heterogeneity of IBD and identify the major transcriptional changes in response to SARS-CoV-2. Extracellular matrix organization (ECMO) was strongly enriched in all organoid groups. Besides being involved in infection processes^85^, ECMO is crucial for the healing response after injury. However, this process can also lead to fibrosis when overstimulated, especially when auxiliary signalling pathways like TGF-β, EGFR and NOTCH, are co-activated^85^, which was the case in CD organoids. In COVID- 19, lung fibrosis emerges after acute lung damage and represents a severe functional impairment and also fibrosis of the kidneys are reported in COVID-19^86, 87^. In parallel, intestinal fibrosis is a feared complication of IBD, especially in CD, appearing as a consequence of chronic inflammation^88^. Therefore, SARS-CoV-2 infection of epithelia might initiate the development of fibrosis in the GI tract, which might aggravate IBD sequels.

Inflammatory signalling pathways were also induced upon infection including interleukin-, TNF-and neurotrophin signalling. The last pathway was strongly induced in infected CD ileum. Besides activating pain, neurotrophins strengthen the epithelial barrier during viral infections, reduce apoptosis and induce TGF-β signalling^89, 90^. Neurotrophin signalling is reported to be induced in IBD^91, 92^, and it is likely that its activation might be implicated in the development of intestinal pain recognized in a substantial fraction of COVID-19 patients. Interestingly, chemokine signalling was only induced in non-IBD organoids, in contrast to interferon signalling, which was induced in h-ileum and UC colon upon infection. Similarly, apoptosis, necroptosis and pyroptosis were enriched only in h-ileum and UC colon. SARS-CoV-2 induces a carefully calibrated caspase-8 activation, leading to the activation of cell-death pathways, which support IL-1β secretion^93^. Necroptosis, apoptosis and *IL1B* expression were strongly upregulated in UC colon upon infection despite the lowest viral load in this organoid group. This reaction was not evident in CD organoids. Recently, a specific subgroup of IBD patients were described, wherein IL-1β-driven neutrophil recruitment perpetuates chronic inflammation in UC. These patients lack responsiveness to anti-TNF therapy^71^ but IL-1β blockage can efficiently suppress inflammation in a murine model^91^. Future research should clarify whether UC patients might generally overreact upon SARS-CoV-2 infection compared to CD patients^7, 74^ and whether this is driven by IL-1β. SARS-CoV-2 induces an interferon response in intestinal organoids that is stronger than from SARS-CoV^25, 95, 96^ and comparable to other intestinal viruses like rotavirus, enterovirus and norovirus^97–99^. Considering interferons themselves, SARS-CoV-2 infection induces low amounts of type I and III interferon transcripts in intestinal organoids^25, 84, 95, 96^. Interferon transcripts were not detectable in our experiment, only their receptors. It was shown that interferons are being actively suppressed by SARS-CoV-2^100^.

By comparing uninfected IBD to non-IBD organoids, we were able to discern the transcriptional changes of the genes involved in the host response to virus already present in IBD. CD ileum organoids showed an induction of several antiviral response genes implicated in antigen presentation (e.g., *HLA-B*), direct antiviral activity and recognition (e.g., *OAS1/L*), cell death (e.g., *FADD*) as well as induction of cyto-and chemokines (e.g., *TNF*). In UC, there was also an induction of direct antiviral activity and recognition (e.g., *MX2, IFIT1*) and important initiators of defence mechanisms (e.g., *IRF7*), but in contrast to CD, response to virus-genes were more often decreased. Together with a reduced expression of SARS-CoV-2 entry factors, CD phenotype likely has a benefit and counteracts infection and viral propagation in ileal epithelial cells. Additionally, CD organoids were generally less reactive in response to the virus compared to UC. In UC, a strong induction of cell death pathways and *IL1*β expression might suppress the virus, but can also lead to increased epithelial stress. Recently, viral persistence in the GI tract was reported in IBD and linked to postacute-COVID-19 symptoms^23^. Whether the transcriptional reaction of CD and UC identified in our model is implicated in the development of long-term sequels, such as the post-acute COVID-19 syndrome, warrants further investigations.

A limitation of our study is still a restricted number of patients from which the organoids were cultivated (N = 5-6), although our study represents a larger cohort compared to other organoid studies using up to three individuals for modelling SARS-CoV-2 infection^25, 35^. Clearly, more patients would be necessary to capture the entire genetic and epigenetic landscape prevalent in IBD. Different infection doses and kinetics might also impact the epithelial transcriptional response in the GI tract. Remitting seeding from the respiratory tract or reinfection from higher GI locations might contribute to COVID-19 GI pathology *in-vivo*. IBD pathology is significantly influenced by immune cells, the connective tissue, and IBD medication, which were not included in our investigations. Nevertheless, our model proved feasible to analyse virus-epithelial interactions of a newly emerging pathogen and to assess GI susceptibility in health and IBD. The model system of intestinal organoids represents intact epithelial tissue with no additional inflammatory stimuli but with the genetic and epigenetic repertoire prevalent in IBD. The fact that infection with SARS-CoV-2 did not lead to higher viral burden in IBD organoids falls in line with the current clinical observation that IBD patients do not seem to be more prone to SARS-CoV-2 infection. Still, we observed differential responses to the virus in IBD organoids such as a stronger activation of neurotrophin and fibrosis-related pathways in CD ileum, and interferon signalling and regulated cell death in UC colon.

Our study indicates that intestinal epithelial cells in IBD have a heightened antiviral state due to persisting changes in gene expression. Together with reductions of *ACE2* and *CTSL/B* levels in CD ileum, this could protect against SARS-CoV-2 infection. On the other hand, as we have seen in UC colon marked with the lowest viral load, it could lead to a possibly detrimental overreaction of the IBD epithelium. These findings should prompt further investigations whether SARS-CoV-2 or other enteric viral infection differently favour stronger GI symptoms, IBD complications or long-term sequels in IBD subtypes.

## Data Availability

The RNAseq data have been deposited in NCBI-GEO under the accession number GSE208684. There are restrictions to the availability of human intestinal organoids generated in this study due to difficulties in their long-term preservation. Further information and requests for resources and reagents should be directed to and will be fulfilled by the lead contact Gregor Gorkiewicz.

## Acknowledgments

We are grateful for the technical support from Martina Loibner, Julia Rieger and Iris Kreuzmann. The valuable contributions from the BSL-3 and the routine laboratories of Institute of Pathology and the Core Facility for Imaging at the Center for Medical Research at Medical University of Graz, as well as the Division of Internal Medicine of the Medical University of Innsbruck are highly acknowledged. G.G. received funding from the Austrian Science fund (FWF, DK-MOLIN W1241) and A.M. is supported by the Christian Doppler research foundation.

## Author contributions

Conceptualization, G.G., A.M., B.J.; Methodology, G.G., A.M., B.J., P.W.; Formal Analysis, B.J., S.B., P.W.; Investigation, B.J., S.B., P.W., N.P., C.W., S.W., M.A., S.E., G.G.; Resources, G.G., A.M., B.T., K.Z.; Data Curation, B.J., S.B.; Writing – Original Draft, B.J., G.G.; Writing – Review & Editing, K.Z., A.M., P.W.,, B.J., G.G.; Visualization, B.J., P.W., S.B.; Supervision, G.G., A.M., Funding Acquisition, G.G., A.M., B.T.,K.Z.

## Materials and Methods

### Ethics approval

Generation and use of organoids were approved by the ethics committee of Medical University Innsbruck, Austria (ethics vote no.: AN4994 323/4.4). All patients gave their informed written consent for participation in this study. SARS-CoV-2 isolation was done during an autopsy study^21^ approved by the ethics committee of the Medical University of Graz (EK-number: 32–362 ex 19/20).

### Human intestinal organoid culture

Intestinal organoids were cultured from biopsy specimens retrieved during endoscopy of IBD patients and healthy controls at the University Hospital Innsbruck. 11 IBD patients and 11 controls were included. Their characteristics are summarized in *Table S1*. Organoid generation was performed with IntestiCult Organoid Growth Medium (STEMCELL Technologies Inc., Vancouver, Canada) with an adapted protocol. Two to three ileal or colonic biopsies, taken from an inflamed site in case of IBD patients, were flushed with ice cold PBS and minced into small pieces with a sterile blade. Tissue was then transferred to 15-ml tubes with 5 ml of Gentle Cell Dissociation Reagent (GCDR, STEMCELL Technologies) and incubated at 4 °C on a rocking platform for 30 min. After centrifugation (290 x g, 5 min, 4 °C), the tissue suspension was transferred to 1 ml of ice cold DMEM (Gibco, Thermo Fisher Scientific Inc., Waltham, Massachusetts, US) with 1% (vol/vol) bovine serum albumin (Roche, Germany). The suspension was gently mixed and passed through a 70 μm cell strainer (Corning, Merck, KGaA, Darmstadt, Germany). The cell suspensions containing whole intestinal crypts were seeded in 50 μl Growth Factor Reduced Matrigel matrix (Corning Inc., Corning, New York, US) on a pre-warmed 24-well plate and allowed to solidify for 10 min at 37 °C, after which 500 μl IntestiCult Growth Medium supplemented with 100 U/ml penicillin and 100 μg/ml streptomycin (Biochrom, Berlin, Germany) was added. Medium was exchanged every two days and organoids were passaged every 5–10 days depending on their growth, with a maximum of 12 passages. During passaging, ileal organoids could be disrupted by pipetting only, as opposed to colonic organoids that needed chemical disruption with Gentle Cell Dissociation Reagent (GCDR) according to the manufacturer’s protocol (STEMCELL Technologies). To create monolayers, organoids were harvested from 24 well plates and processed according to protocol (STEMCELL Technologies); in brief, they were mechanically disrupted by pipetting, trypsinized, washed and seeded in a 96 well plates pre-coated with 5% Matrigel. Four monolayers were created for each patient in order to have two replicates for infection and two for control. Remaining 3D organoids were passaged further and split into two wells for each patient, one for each treatment.

### Virus isolation and culture

The used SARS-CoV-2 strain originated from an autopsy study performed at the first pandemic peak^24^. The strain hCoV-19/Austria/Graz-MUG5 (B.1.5/O/20A) originated from a lung swab from a patient with concomitant high-level intestinal SARS-CoV-2 carriage. The virus strain was propagated using Vero CCL-81 cells^24^. To prepare the inoculum for organoid infections, Vero CCL-81 cell supernatants were centrifuged at 1500 rcf for 10 min at RT and SARS-CoV-2 particles were quantified by using a qPCR as described^24^.

### Organoid infection

Organoid infection followed protocols adapted from Lamers *et al.*^25^ and Ettayebi *et al.*^26^. Briefly, the viral inoculum was prepared by diluting the virus corresponding to 10^5^ PFU in serum-free (AD) medium containing Advanced DMEM/F-12 (Thermo Fisher Scientific Inc.), 1M HEPES (Merck), 1% GlutaMAX (Thermo Fisher Scientific) and 1% Pen/Strep (Thermo Fisher Scientific). Conditioned medium from Vero CLL-81 cells without virus was used as control for uninfected (mock) samples. Monolayers in 96-well plates were infected on day 13 after seeding. They were washed with 150 µl of AD medium ahead of the infection and then treated with 100 µl of inoculum or mock medium. 3D organoids were washed with 500 µl of AD medium and left for 5 minutes for the washing medium to penetrate the Matrigel. Upon removal of AD medium, 500 µl of the inoculum or mock medium was added to the organoids. Infection dose corresponded to a multiplicity of infection (MOI) of 0.1 for monolayers and a MOI of 1 for 3D organoids. Monolayers and 3D organoids were incubated in an atmosphere containing 5% CO_2_ at 37 °C for 2 h to allow for the virus to attach. Supernatants were then removed and the cells were washed twice with 200 µl AD medium for 5 minutes. 150 µl and 500 µl of IntestiCult™ medium was added to monolayers and organoids, respectively, and incubated for further 48 hrs. Thereafter, 200 µl of TRIzol (Thermo Fisher Scientific) was added to each monolayer well and replicates were collected separately. Cell lysates in TRIzol were vortexed once and immediately frozen at −20 °C until further processing.

### Immunofluorescence

72 hours post infection 3D organoids embedded in Matrigel were fixed in Carnoy’s solution, embedded in paraffin, sectioned and mounted on glass slides. After deparaffinization with Xylol (SAV Liquid Production, Germany) and rehydration in a series of ethanol dilutions (Supelco, Germany; 100%, 90%, 70%, 50%), samples were blocked with 3% normal goat serum and 1% bovine serum albumin (Roche) in PBS (Gatt-Koller, Austria) and 0.05% Tween20 (PBST; Roche, Germany) at room temperature for 1 hour. Samples were incubated overnight at 4 °C with a rabbit polyclonal antibody against the SARS-CoV-2 spike protein (40150-T62-COV2; Sino Biological, Germany) diluted 1:500 in PBST (Roche, Germany) with 1% goat serum. After washing in PBST, samples were incubated for 1 hour at room temperature with the goat anti-rabbit secondary antibody Alexa Fluor™ 488 (A11034; Thermo Fisher Scientific, US) diluted 1:1000 in PBST with 1% goat serum. Images were captured with a Nikon A1R HD25 confocal microscope and NIS-Elements software was used for image acquisition and processing.

### RNA isolation and sequencing

Total RNA was isolated with TRIzol (Thermo Fisher Scientific) according to manufacturer’s protocol. RNA quality was controlled by using a Bioanalyzer instrument (Agilent) and sequencing libraries were prepared from 25-250ng of total RNA per sample following Roche’s strand specific “KAPA RNA Hyper Prep Kit + RiboErase (HMR)” library preparation protocol for double indexed Illumina libraries. Breifly, the rRNA fraction was depleted using complementary DNA oligonucleotides; rRNA-DNA hybrids and DNA baits were removed by treatment with RNase H and DNase. RNA was heat-fragmented and subjected to first strand synthesis using random priming. The second strand was synthesized incorporating dUTP instead of dTTP to preserve strand information. After A-tailing Illumina sequencing compatible adapters carrying unique dual indices were ligated (NEXTFLEX® Unique Dual Index Barcodes). Following bead-based clean-up steps the libraries were amplified using 13 cycles of PCR. Library quality and size was checked with the qBit, Bioanalyzer (Agilent) and qPCR. Sequencing was carried out on an Illumina NovaSeq6000 system in PE100bp mode yielding between 20-40 million fragments per sample. Following base calling, adaptor clipping was performed using cutadapt 2.4^101^. Data was mapped against the hg38.p12 genome including the SARS-CoV-2 genome (A9351 strain) using STAR v2.7.5a^102^. Gene-wise read count tables were used for analysis with DESeq2^103^ and transformed to visualize sample distances by principal component analysis (PCA).

### Viral load

Viral gene counts were normalized by the R package DESeq2^103^, summed by sample and log_2_ transformed. To compare the groups regarding the viral load, a mean of monolayer replicates was used resulting in a single value per patient. Compared groups were healthy ileum vs. healthy colon, healthy ileum vs. CD ileum, and healthy colon vs. UC colon.

### Differential gene expression analysis

Differences in gene expression were analysed using the R package DESeq2^103^. Two monolayer replicates from a certain patient and treatment condition were pooled during the analysis after confirming a high correlation (Suppl.Fig.1). The effects of SARS-CoV-2 infection were determined for each group separately and by pairing the infected with the control sample originating from the same patient. If uninfected or infected sample was missing due to low sequencing quality, this patient was removed from the subsequent analyses resulting in the final number of included patients: 6 h-ileum, 5 CD ileum, 5 h-colon and 5 UC colon. When assessing the differences in gene expression due to IBD itself, control samples from an IBD group were compared to control samples from the corresponding healthy organ resulting in the final number of included patients: 6 h-ileum, 6 CD ileum, 5 h-colon and 5 UC colon.

### Pathway analyses

For each group, two types of functional pathway analyses were performed using the R package clusterProfiler^104^ and the Reactome pathway database^41^. Over-enrichment analysis (ORA) was based on a list of DEGs upon SARS-CoV-2 infection with *P* < 0.05. Gene set enrichment analysis^105^ (GSEA) was based on all expressed genes pre-ranked by their *P* value with the sign of LFC. The rank metric was calculated by the following formula:

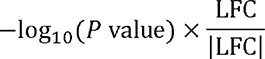

### Statistical analysis

GraphPad Prism and R were used for data analysis and imaging. All data are represented as mean ± SD if not otherwise specified. Statistics of differential gene expression were calculated with the Wald test. Other statistical significance testing employed the Mann-Whitney-U-Test and 1way ANOVA with uncorrected Fisher’s LSD. Correlation analyses employed Pearson correlation. *P* values < 0.05 were considered statistically significant. Benjamini-Hochberg (BH) procedure with a False Discovery Rate (FDR) < 0.1 was used to determine the adjusted *P* values (*Padj*) in the differential gene expression analysis. Statistical tests are also described within the figure legends.

## Supplemental information legends

**Table S1. Characteristics of healthy controls and IBD patients for generation of intestinal organoids**

**Supplementary Figure 1** – **Correlation of monolayer replicates.** Each plot shows correlation of normalized gene counts of two organoid-derived monolayer replicates from the same patient. Red dots represent genes aligned to SARS-CoV-2 genome and black represent normalized human reads.

**Supplementary Figure 2 – Characteristics of uninfected and infected organoids. A)** Representative immunofluorescence staining of the SARS-CoV-2 spike (S) protein (green) in uninfected 3D organoids. DAPI counterstain (blue). Bar represents 50 μm. **B)** Principal component analysis of h-ileum, CD ileum, h-colon and UC colon, infected with SARS-COV-2 or uninfected. Sample distances are calculated from transformed normalized gene counts and visualized on PC1-PC5 explaining variance percentage as stated. Each dot represents pooled monolayer replicates from one patient, either infected or uninfected. **C)** Expression of mitochondrial genes as a proxy for cell damage is not different between uninfected and SARS-CoV-2 infected healthy and IBD organoids. Dots represent single organoid monolayer replicates. Mean of log(normalized gene counts) with SD; Mann-Whitney-U-Test. N(patients) = 5-6; CD = Crohn’s disease; UC = ulcerative colitis; ile = h-ileum; col = h-colon.

**Supplementary Figure 3 – Expression of *TMPRSS4*, *FURIN* and *NRP1* do not differ between the groups.** Each dot represents pooled organoid monolayer replicates from one patient. Mean of normalized gene counts with SD; 1way ANOVA, uncorrected Fisher’s LSD on following comparisons: h-ileum vs. CD ileum, h-ileum vs. h-colon, h-colon vs. UC colon. N(patients) = 5-6; CD = Crohn’s disease; UC = ulcerative colitis; * *P* < 0.05, ** *P* <0.01, *** *P* < 0.001, **** *P* < 0.0001.

**Supplementary Figure 4** – **Correlation analysis, extended.** Viral load correlates to normalized gene counts of SARS-CoV-2 entry-related genes in samples: 1) all combined, 2) by group, 3) by organ. Each dot represents one patient, pooled organoid monolayer replicates. Viral load is defined as the sum of viral genes. Pearson correlation of transformed normalized gene counts. N(patients) = 5-6; CD = Crohn’s disease; UC = ulcerative colitis.

**Supplementary Figure 5 – Shared and unique DEGs upon infection with SARS-CoV**-**2. A)** Upregulated (up) and downregulated (down) DEGs (*Padj* < 0.1) minimally overlap between healthy and IBD organoids separated by organ. Upregulated and downregulated DEGs with **B)** *P* < 0.05 and **C)** *Padj* < 0.1 overlap more between organoids from colon and ileum of healthy origin as opposed to IBD organoids. Statistics of differential gene expression were calculated with the Wald test. N(patients) = 5-6; CD = Crohn’s disease; UC = ulcerative colitis; IBD = inflammatory bowel disease.

**Supplementary Figure 6 – Enrichment map of gene set enrichment analysis (GSEA, Reactome).** Upregulated pathways are shown in red and downregulated pathways in blue (*Padj* < 0.05). Pathways which share genes contributing to the enrichment score are connected with a line. Size of circles corresponds to the number of contributing genes. N(patients) = 5-6; CD = Crohn’s disease; UC = ulcerative colitis.

**Supplementary Figure 7 – Organoids from IBD patients are transcriptionally different from healthy organoids.** Volcano plots of differential gene expression in uninfected **A)** CD ileum compared to h-ileum and **B)** UC colon compared to h-colon. Dots represent genes; horizontal dashed line marks *P* value of 0.05; vertical dashed lines mark LFC of 0.4. Statistics of differential gene expression were calculated with the Wald test. N(patients) = 5-6; CD = Crohn’s disease; UC = ulcerative colitis; FC = fold change; NS = non-significant.

**Supplementary Figure 8 – Organ and health status influence expression of genes involved in response to virus after SARS-CoV-2 infection of organoids.** Volcano plots of differential gene expression upon infection. Dots represent genes; horizontal dashed line marks *P* value of 0.05; vertical dashed lines mark log_2_ FC of -/+0.6. Statistics of differential gene expression were calculated with the Wald test. N(patients) = 5-6; CD = Crohn’s disease; UC = ulcerative colitis; FC = fold change; GOI = gene of interest.

### Supplementary files

Supplementary File 1, Differential gene expression upon infection

Supplementary File 2, Pathway analysis ORA Reactome

Supplementary File 3, Pathway analysis GSEA Reactome

Supplementary File 4, Differential gene expression IBD

Supplementary File 5, Response to virus gene set analyses

