## Supplementary figures for "Reduced SARS-CoV-2 infection and altered antiviral transcriptional response in IBD intestinal organoids"

**Table S1. Characteristics of healthy controls and IBD patients for generation of organoids**

| Group | | h-ileum | CD ileum | h-colon | UC colon |
|---|---|---|---|---|---|
| **Number of patients** | | 6 | 6 | 5 | 5 |
| **Age in years** | | | | | |
| | mean (SD) | 50.3 (14.3) | 37.2 (14.3) | 45.2 (20.1) | 48.2 (16.3) |
| | range | 26-65 | 18-58 | 20-72 | 21-71 |
| **Female/male** | | 5/1 | 3/3 | 2/3 | 3/2 |

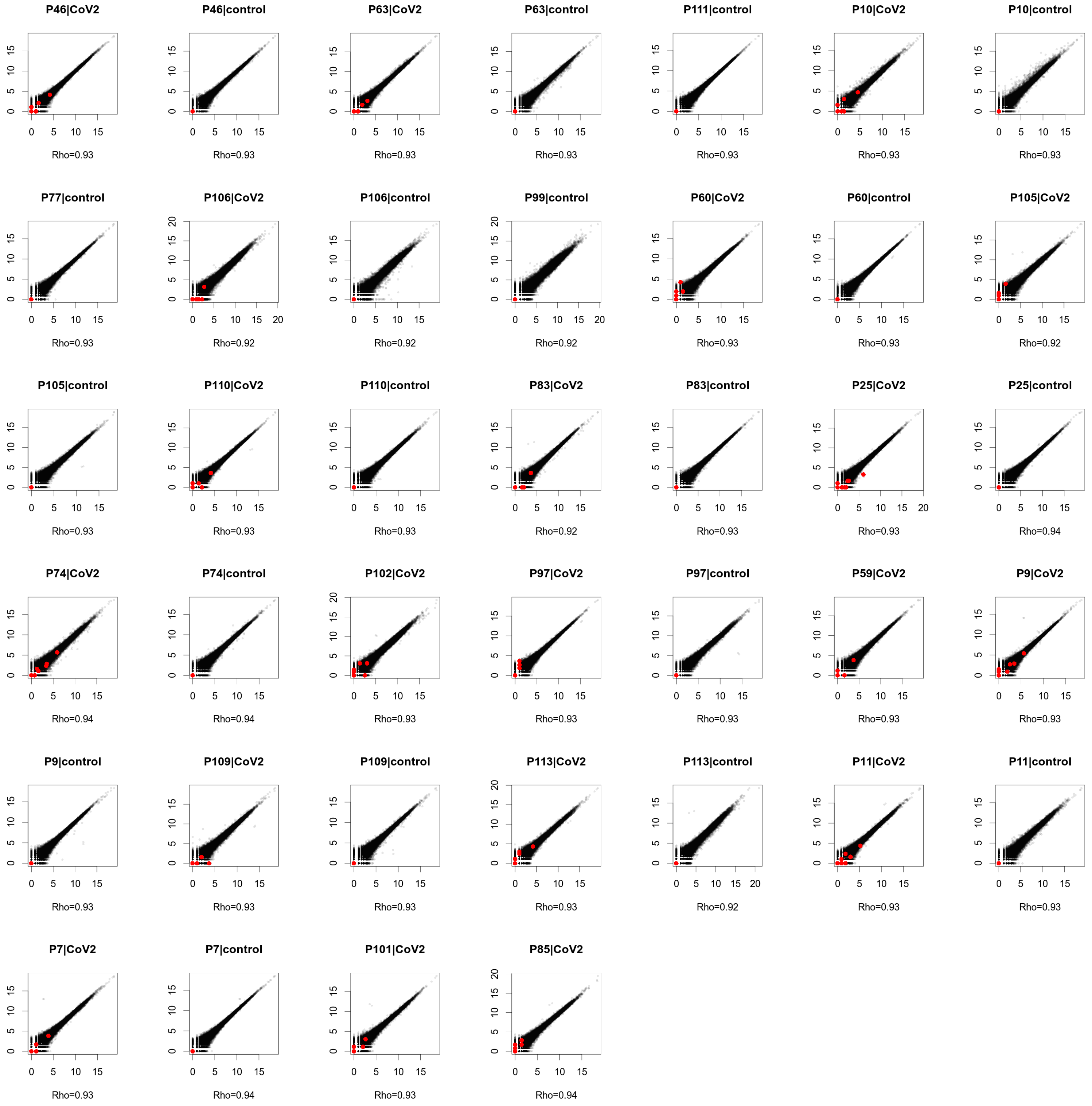


**Supplementary Figure 1 – Correlation of monolayer replicates.** Each plot shows correlation of normalized gene counts of two organoid-derived monolayer replicates from the same patient. Red dots represent genes aligned to SARS-CoV-2 genome and black represent normalized human reads.

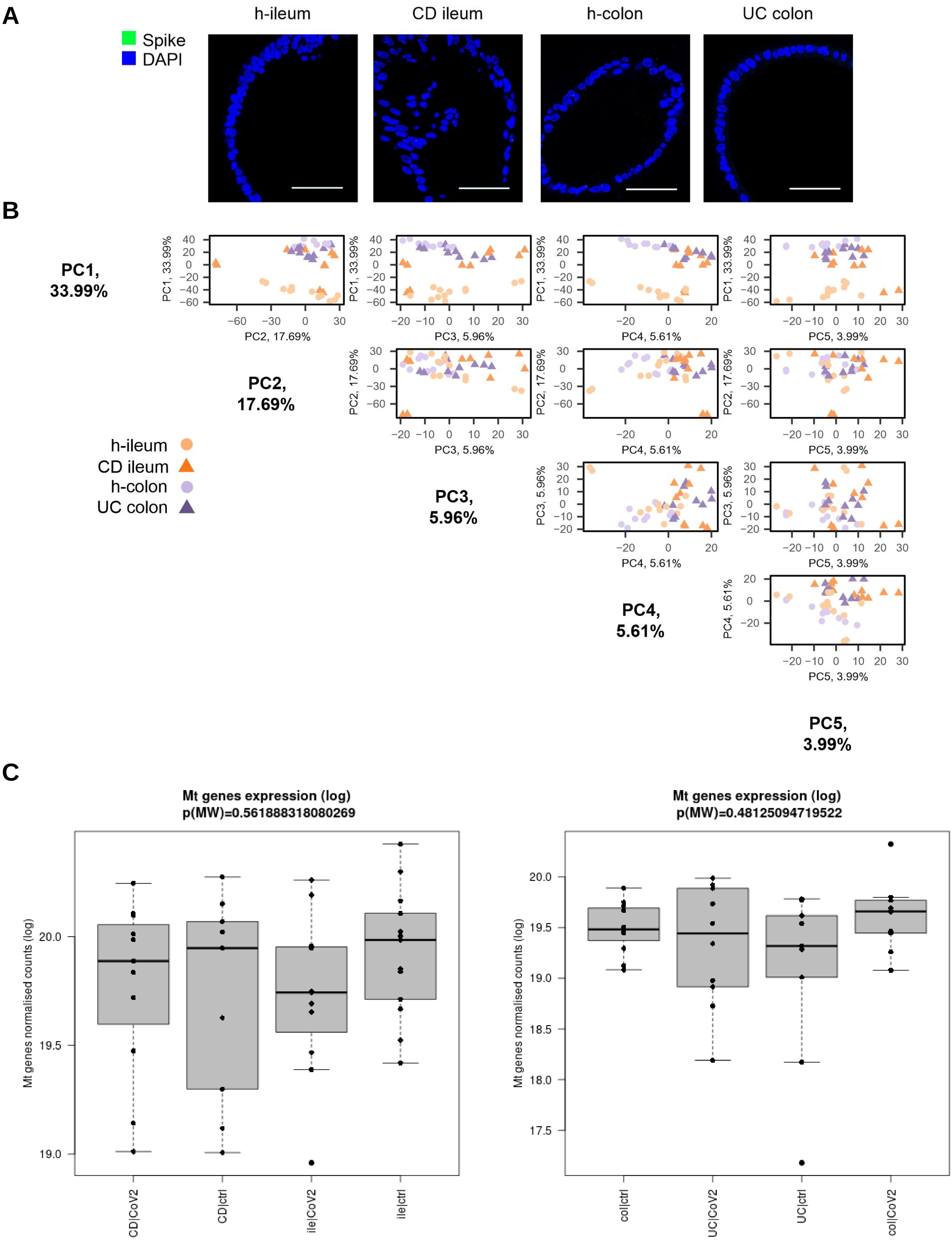


**Supplementary Figure 2 – Characteristics of uninfected and infected organoids. A)** Representative immunofluorescence staining of the SARS-CoV-2 spike (S) protein (green) in uninfected 3D organoids. DAPI counterstain (blue). Bar represents 50 µm. **B)** Principal component analysis of h-ileum, CD ileum, h-colon and UC colon, infected with SARS-COV-2 or uninfected. Sample distances are calculated from transformed normalized gene counts and visualized on PC1-PC5 explaining variance percentage as stated. Each dot represents pooled monolayer replicates from one patient, either infected or uninfected. **C)** Expression of mitochondrial genes as a proxy for cell damage is not different between uninfected and SARS-CoV-2 infected healthy and IBD organoids. Dots represent single organoid monolayer replicates. Mean of log(normalized gene counts) with SD; Mann-Whitney-U-Test. N(patients) = 5-6; CD = Crohn’s disease; UC = ulcerative colitis; ile = h-ileum; col = h-colon.

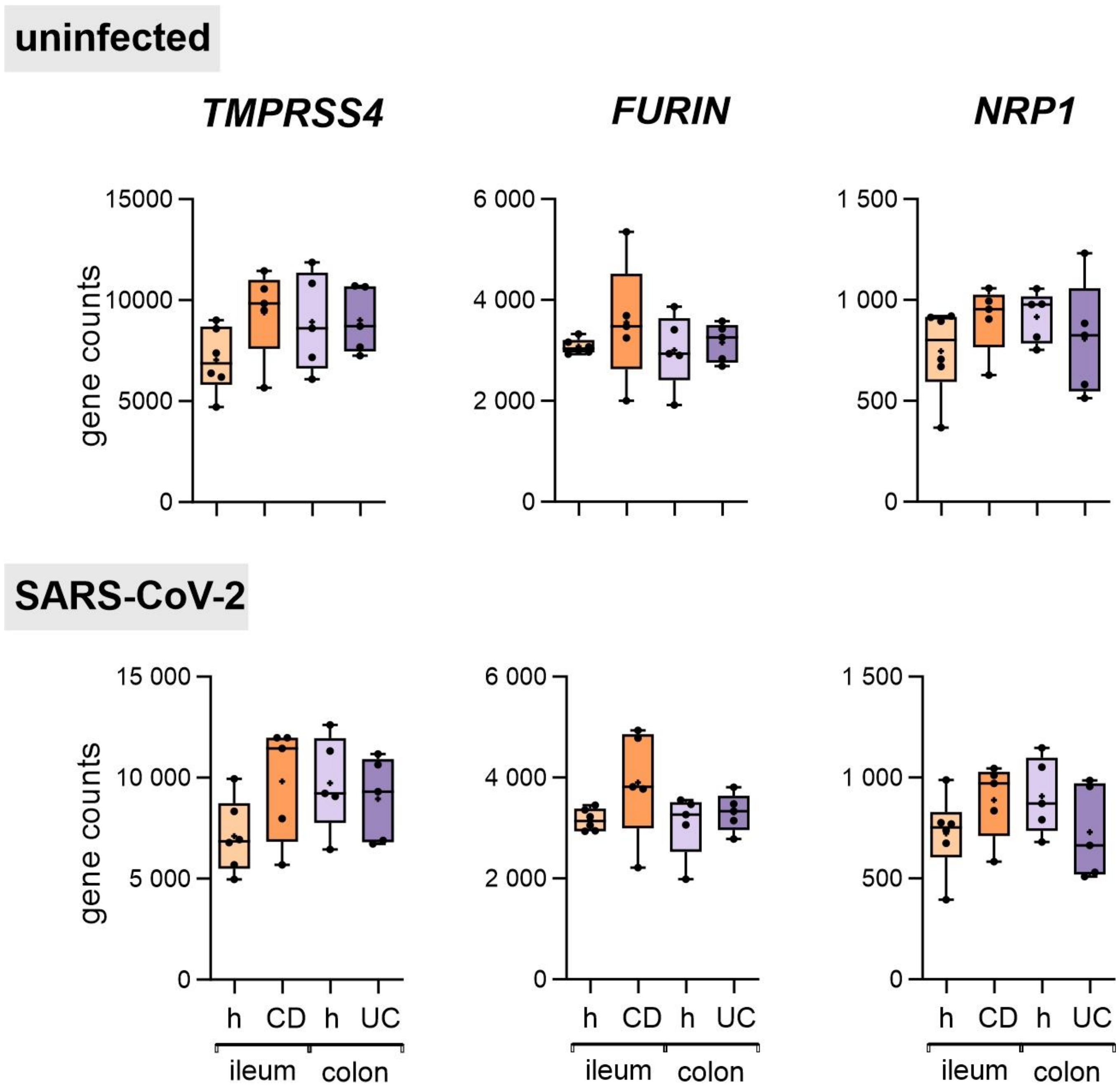


**Supplementary Figure 3 – Expression of *TMPRSS4*, *FURIN* and *NRP1* do not differ between the groups.** Each dot represents pooled organoid monolayer replicates from one patient. Mean of normalized gene counts with SD; 1way ANOVA, uncorrected Fisher's LSD on following comparisons: h-ileum vs. CD ileum, h-ileum vs. h-colon, h-colon vs. UC colon. N(patients) = 5-6; CD = Crohn's disease; UC = ulcerative colitis; * $P < 0.05$, ** $P < 0.01$, *** $P < 0.001$, **** $P < 0.0001$.

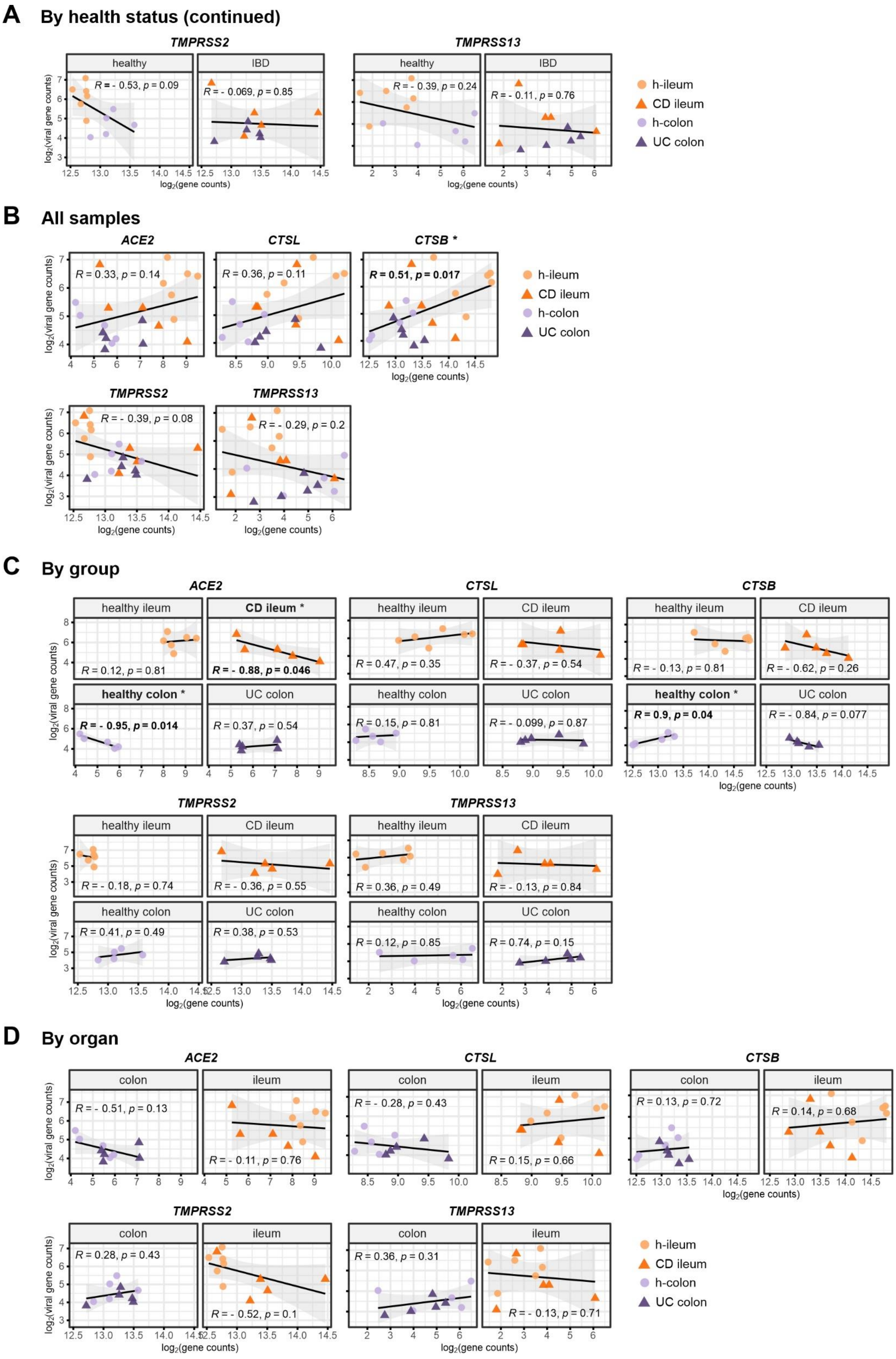


**Supplementary Figure 4 – Correlation analysis, extended.** Viral load correlates to normalized gene counts of SARS-CoV-2 entry-related genes in samples: 1) all combined, 2) by group, 3) by organ. Each dot represents one patient, pooled organoid monolayer replicates. Viral load is defined as the sum of viral genes. Pearson correlation of transformed normalized gene counts. N(patients) = 5-6; CD = Crohn's disease; UC = ulcerative colitis.

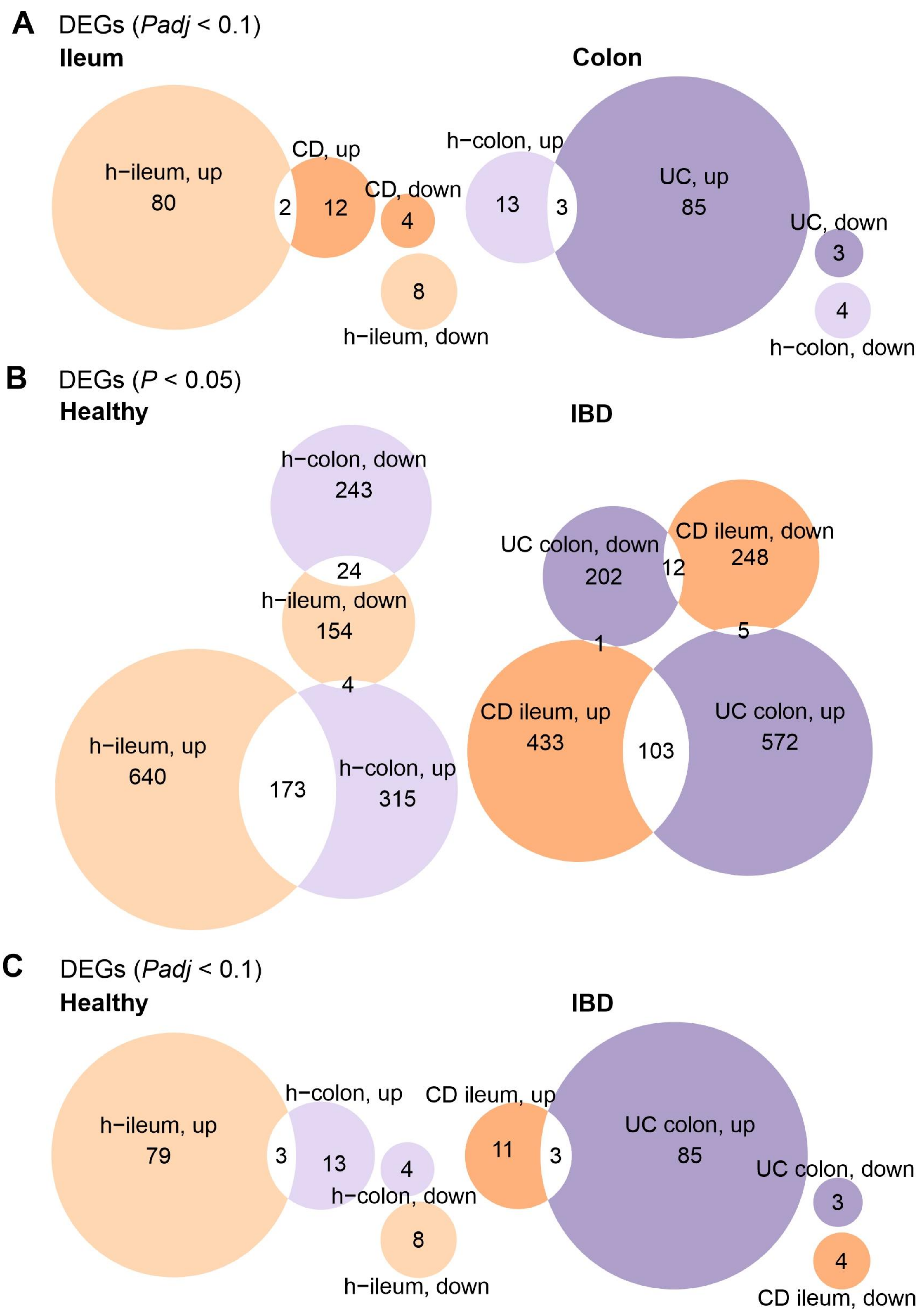


**Supplementary Figure 5 – Shared and unique DEGs upon infection with SARS-CoV-2. A)** Upregulated (up) and downregulated (down) DEGs (*Padj* < 0.1) minimally overlap between healthy and IBD organoids separated by organ. Upregulated and downregulated DEGs with **B)** *P* < 0.05 and **C)** *Padj* < 0.1 overlap more between organoids from colon and ileum of healthy origin as opposed to IBD organoids. Statistics of differential gene expression were calculated with the Wald test. N(patients) = 5-6; CD = Crohn’s disease; UC = ulcerative colitis; IBD = inflammatory bowel disease.

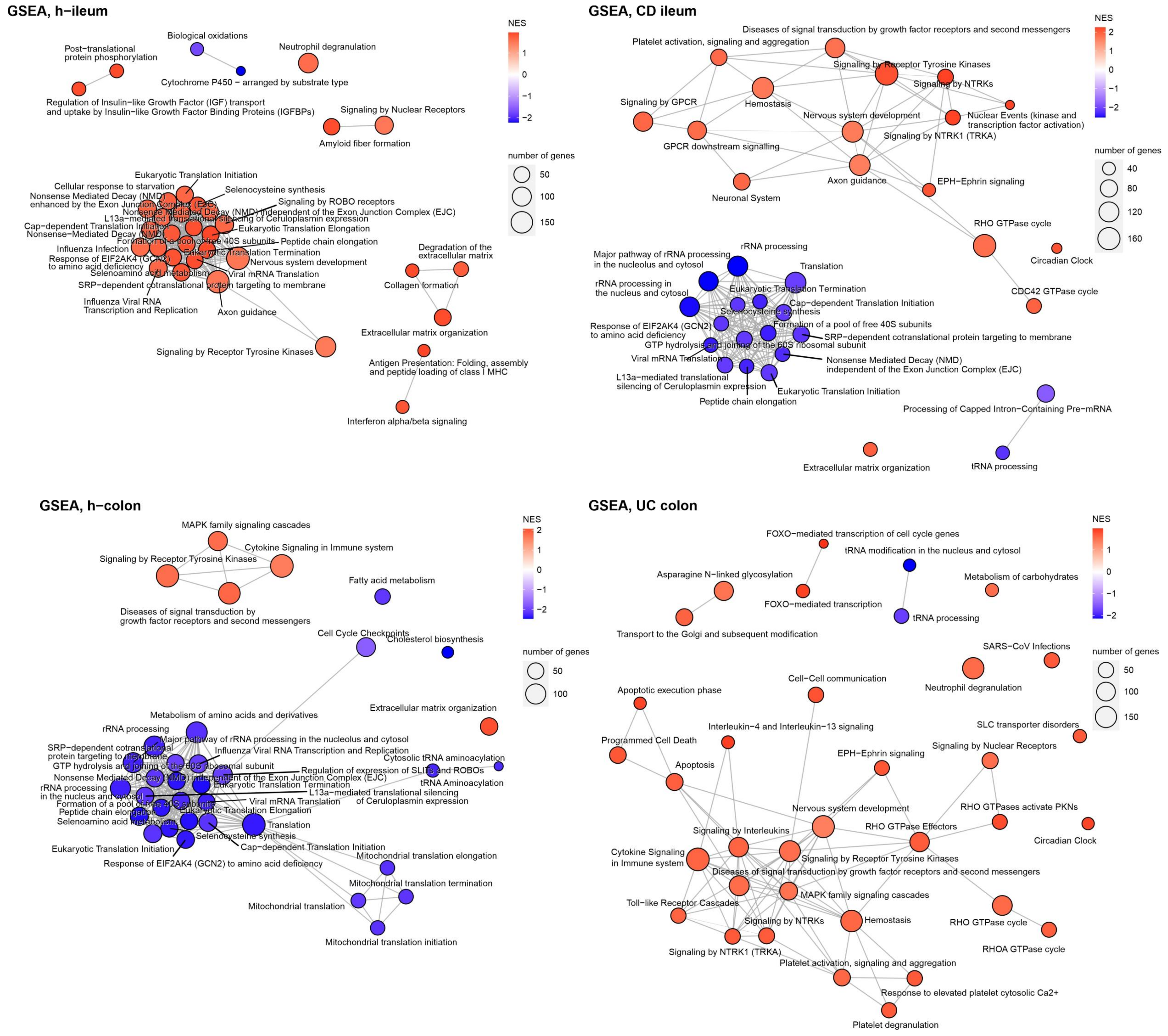


**Supplementary Figure 6 – Enrichment map of gene set enrichment analysis (GSEA, Reactome).** Upregulated pathways are shown in red and downregulated pathways in blue (*Padj* < 0.05). Pathways which share genes contributing to the enrichment score are connected with a line. Size of circles corresponds to the number of contributing genes. N(patients) = 5-6; CD = Crohn's disease; UC = ulcerative colitis.

**A**

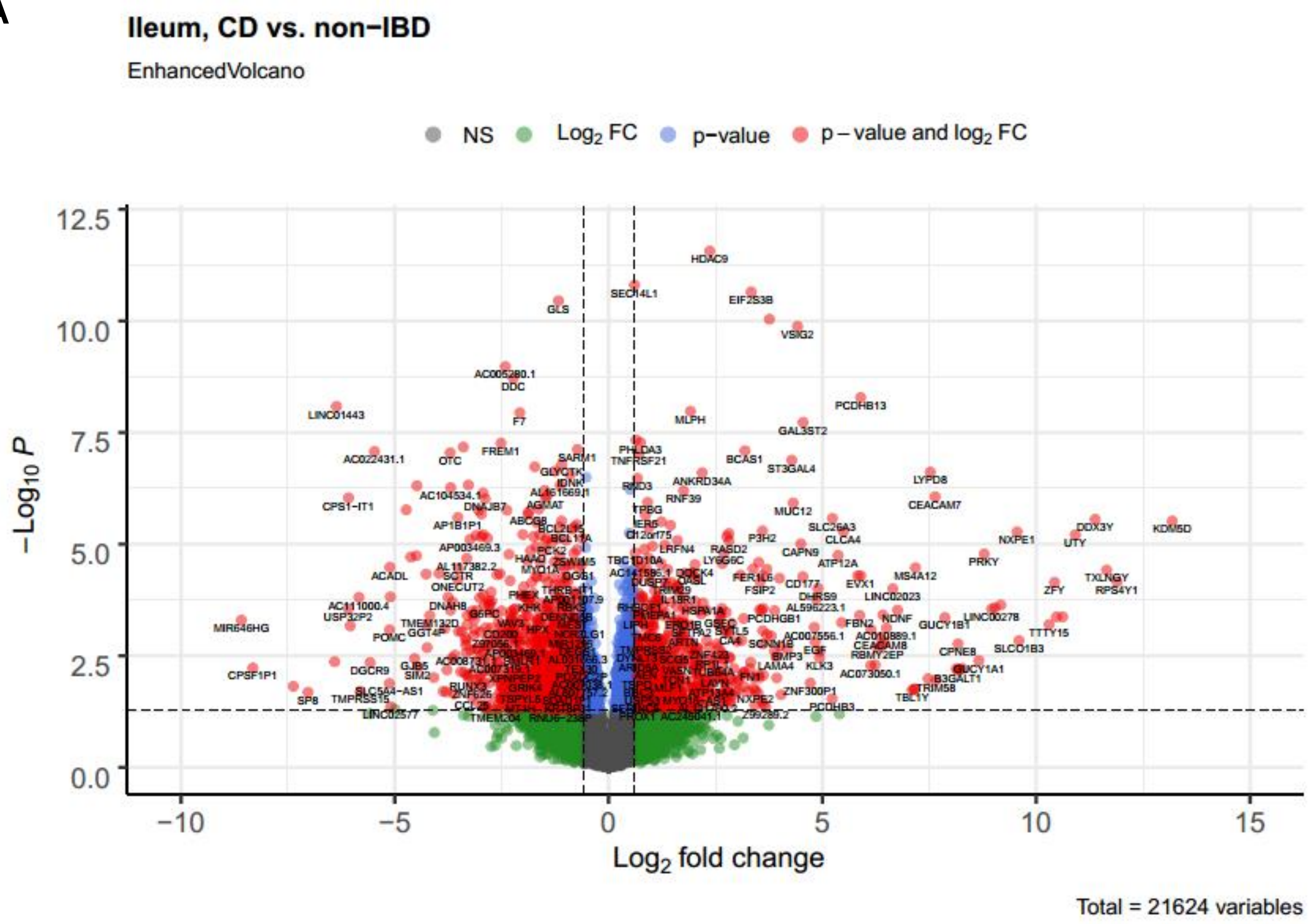


**B**

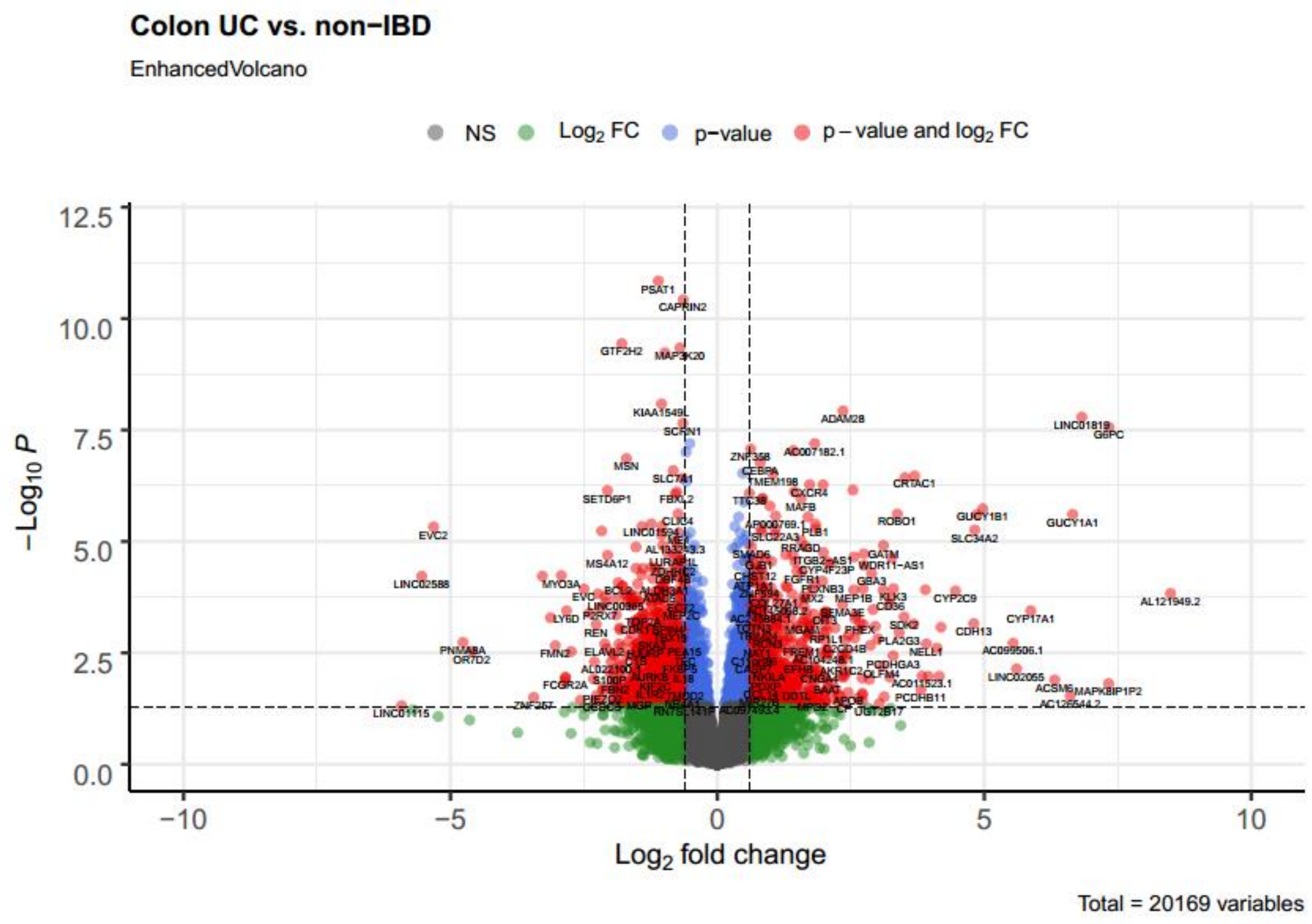


**Supplementary Figure 7 – Organoids from IBD patients are transcriptionally different from healthy organoids.** Volcano plots of differential gene expression in uninfected **A)** CD ileum compared to h-ileum and **B)** UC colon compared to h-colon. Dots represent genes; horizontal dashed line marks *P* value of 0.05; vertical dashed lines mark LFC of 0.4. Statistics of differential gene expression were calculated with the Wald test. N(patients) = 5-6; CD = Crohn’s disease; UC = ulcerative colitis; FC = fold change; NS = non-significant.

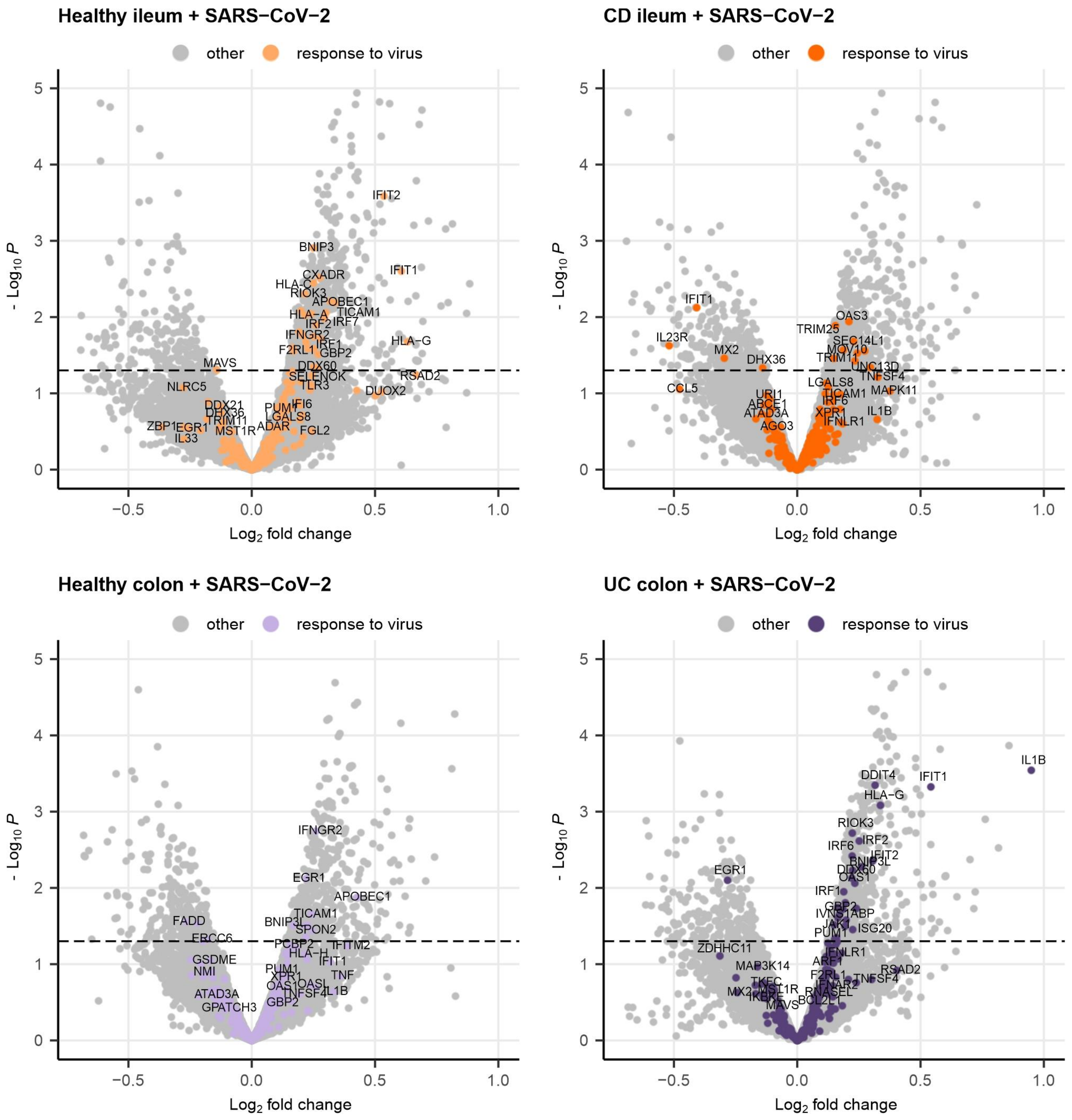


**Supplementary Figure 8 – Organ and health status influence expression of genes involved in response to virus after SARS-CoV-2 infection of organoids.** Volcano plots of differential gene expression upon infection. Dots represent genes; horizontal dashed line marks *P* value of 0.05; vertical dashed lines mark $\log_2$ FC of -/+0.6. Statistics of differential gene expression were calculated with the Wald test. N(patients) = 5-6; CD = Crohn's disease; UC = ulcerative colitis; FC = fold change; GOI = gene of interest.
